# Bayesian Spatiotemporal Projection of Chagas Disease Incidence in Brazil

**DOI:** 10.1101/2023.08.29.23294788

**Authors:** Ethan Roubenoff

## Abstract

Chagas Disease is a parasitic infection caused by the *T. Cruzi* parasite endemic to Central and South America and transmitted through contact with *Triatomine* insects, commonly known as “kissing bugs.” Although the symptoms of Acute Chagas Disease (ACD) are nonspecific, untreated chronic infection can lead to heart disease, enlarged esophagus and colon, and stroke. Chagas disease has become increasingly rare owing to a series of public health interventions, including insect eradication campaigns in Brazil through the 1980’s that considerably reduced the number of new acute cases. However, hundreds of new acute cases still are diagnosed annually, primarily in the states of Pará, Amapá, and Acre. Moreover, the population in areas of high Chagas endemicity are changing: many areas are growing and becoming increasingly urban, whereas others are decreasing in population. We estimate the Incidence Rate (IR) for Acute Chagas disease over the period 2001-2019 in Brazil at the municipal level and investigate the variation of these rates with climatic factors. These estimates are used to project forward incidence of Acute Chagas Disease over the following decade 2020-2029. Modeling ACD presents numerous methodological challenges since incidence is rare, with extreme overdispersion of zero-case counts, and vectors exhibit a highly spatially- and temporally-clustered pattern. We use a spatially- and temporally-autoregressive small-area smoothing models to estimate the true latent risk in developing Acute Chagas Disease. The Bayesian model presented here involves spatio-temporal smoothing via a Zero-Inflated (Lambert 1992), Knorr-Held (2000)-Type spatio-temporal model with a BYM2 (Morris, 2019) spatial convolution to predict smoothed incidence rates of Chagas disease. As well, we include estimates of Brazil’s growing population and projected bioclimate to evaluate how climate and population change may affect ACD rates. We estimate that cases will continue to increase in the absence of control efforts, primarily driven by a growing peri-urban population in regions of Chagas endemicity.

## 1 Introduction

Chagas disease is a vector-borne parasitic infection in humans caused by the *T. Cruzi* parasite, and is transmitted to humans primarily through contact with infected *Triatomine* insects commonly known as “kissing bugs” (WHO Expert Committee on the Control of Chagas Disease 2002; Pérez-Molina and Molina 2018; Canals et al. 2017). Transmission of *T. Cruzi* to humans can occur following bites from infected kissing bugs or contact with their feces; human-human transmission can occur via blood transfusion and congenitally from mothers to children via the vertical pathway. Over 80% of transmission occurs from human-vector contact, with congenital transmission responsible for nearly all other new infections; screening of blood donation has nearly eliminated all transmission from transfusions (World Health Organization 2015). Acute symptoms of Chagas disease include fever, inflammation of the infection site, eyelid edema, and swollen lymph nodes and tonsils. Acute symptoms resolve spontaneously over 4-8 weeks and treatment during the acute phase with antiparasitic medication is highly effective at curing infection (Bern et al. 2007). However, since acute symptoms are generally nonspecific and the burden of infection affects many communities lacking affordable access to high-quality healthcare, many acute cases go undiagnosed and untreated. Untreated chronic Chagas infection can cause cardiomypoathy, megacolon, stroke, and megaesophagus in 30-40% of patients in the 10-30 years following acute infection (Pérez-Molina and Molina 2018; Sosa-Estani and Segura 2015). Although Chagas is fairly rare—acute infection incidence is on the order of a few hundred infections annually in Brazil for a population of around 200 million—it is this possibility of chronic complications, especially cardiomyopathy, that makes early identification and control of Chagas an important public health concern.

Chagas disease is also known as a disease of poverty affecting mostly poor and indigenous rural communities in South America (Fernández, Gaspe, and Gürtler 2019; Sosa-Estani and Segura 2015; Dias 1987; Tarleton et al. 2007). Many people are at increased risk of infection due to the use of certain residential construction materials hospitable to *Triatomine* infestation, especially untreated wood. People working certain jobs that entail contact with *Triatomine* habitats—including forestry or agriculture—may be at increased risk of exposure during employment. A number of non-pharmaceutical interventions can alleviate much of the probability of contact, including insecticide usage, bed netting, and removal of certain residential construction materials known to be *Triatomine* habitats. Intervention campaigns through the 1980’s focused directly on the class dynamics of Chagas risk by implementing control efforts across the sociodemographic ladder (Dias 1987), yet many persisting Chagas hotspots occur in poor and rural parts of Brazil. Controlling the incidence of Chagas remains an important issue of equity.

Since then, Public health efforts to eliminate new Chagas infections and pharmaceutical developments to treat latent chronic infections and complications have been successful at reducing the burden of Chagas disease in Brazil (World Health Organization 2015; Sosa-Estani and Segura 2015). The two primary methods of transmission—human-vector contact, either through bites or contact with vector feces, and the vertical pathway from mothers to infants—have have required vastly different interventions, both with success. Early studies dating to the late 1940s found that continuous use of residential insecticides was highly effective at eliminating transmission, indicating that residential contact with *Triatomines* may have been responsible for the majority of transmission risk (Dias 1987). Eradication programs by Brazil’s SUCAM (Superintendencia de Campanhas de Saude Publica) in the 1980s involved identifying areas of risk by sampling insects in homes and generating maps of high risk locations. All homes within more than 700 high-risk municipalities, regardless of known infestation, were sprayed with insecticide every 3-6 months until under 5% of homes were found to be infested with any insects and no *Triatomines* were found in any homes. Overall, more than 5 million homes were sprayed with insecticide, resulting in a 73% reduction in the number of infested homes by 1986 (*ibid*.) and the total elimination of transmission by *T. infestans*—previously the vector responsible for most transmission—resulting in a 94% reduction in new acute cases (Gurgel-Gonçalves et al. 2012).

Congenitally-transmitted Chagas disease via the vertical pathway is less frequent than transmission via contact with *Triatomines* (World Health Organization 2015). Screening for Chagas Disease among potential mothers living in high-risk areas and initiating treatment in advance of pregnancy is ideal for reducing the probability of successful vertical transmission, although commencing treatment after pregnancy appears to be well tolerated by both the mother and the fetus (Cevallos and Hernández 2014). While it is not currently possible to entirely eliminate vertical transmission, treatment of infants with suspected Chagas infection within the first year of life is very successful at eliminating the disease from children (Carlier et al. 2011; Moya, Basso, and Moretti 2005). Transmission may occur at any time during pregnancy, but is theorized to be more likely to occur during the second and third trimesters (*ibid*). Most congenital transmission occurs from mothers who are in the chronic phase of disease, however vertical transmission has been documented from mothers who are acutely infected at conception or become infected during the course of pregnancy. It has been proposed that the level of parasitemia of the mother may affect the probability of vertical transmission and the severity of infection at birth (Carlier et al. 2011).

Antiparasitic medications benzniazole and nifurtimox have proven efficacy against Chagas disease, and the former is usually recommended for treatment (Bern et al. 2007). If treated in the acute phase, complete parasitological cure can occur in 60-85% of vector-transmitted infections and more than 90% of congenital infections when treatment is administered within the first year of life (Altcheh et al. 2011; Carlier et al. 2011; Cevallos and Hernández 2014; Moya, Basso, and Moretti 2005). If Chagas disease is left untreated until the chronic phase, treatment is less effective— only 60% of participants achieved negative serology within 3-4 years. Even if not resulting in a complete cure, treatment may slow the development of Chagas cardiomyopathy and other potentially lethal complications, and treatment is recommended for all patients presenting with positive serology (*ibid*.).

Despite progress towards elimination, there are still an estimated 1-4.6 million people currently infected with chronic Chagas disease and approximately 6,000 deaths per year (Simões et al. 2018). Pérez-Molina and Molina 2018 estimate that in 2010, over 70 million people were at risk of contracting Chagas disease across 21 countries in Latin America, and 38,593 new infections were reported that year. This count of acute infections down considerably from 55,585 in 2005 and from more than 700,000 between 1980-1985. Most individuals living with Chagas disease are located in three countries—Argentina, Brazil, and Mexico—and the most new infections were reported in Bolivia (World Health Organization 2015). While preventative interventions have brought the new infection rate down considerably in Brazil, the rate of decrease has not necessarily been equal across the country. The WHO estimates that as of 2015, the incidence of new Chagas infections in Brazil via human-vector contact was 0.084 per 100,000 population and via congential transmission 0.020 per 100 live births (*ibid*.). Our analysis aides in identifying areas where future interventions can further alleviate risk of the disease.

The distribution of *triatomines* is a highly spatial process within endemic areas, and as a result risk of contracting Chagas disease is a complex interaction of local vector population, local human population, and interaction between the two. Despite the elimination of *T. infestans*, previously the vector responsible for the most new cases of Chagas Disease, 62 known species of *Triatomines* in Brazil are responsible for transmission; some, including *Panstrongylus geniculatus* and *P. megistus*, are widespread over the country, whereas others are more localized spatially (Gurgel-Gonçalves et al. 2012). Certain biomes, including the Cerrado tropical savannah in the central region and Caatinga shrub forest in the northeast, have a higher diversity of species. Not all species of *Triatomines* are equally likely to transmit Chagas disease to humans;for example, while the most epidemiologically relevant species may be *P. megistus*, which is common in domiciles and a frequent carrier of *T. cruzi*, the behavior and habitat of *T. sordida* is more likely to result in contact from agricultural activities but unlikely to result in residential contact. Gurgel-Gonçalves et al. 2012 remark that nearly all areas of Brazil have some risk of Chagas disease, but certain regions, especially the Cerrado and Caatinga, present more risk.

Climate change presents an ambiguous threat to incidence of Chagas disease. Tamayo et al. 2018 find that *Triatomine* vectors of Chagas disease may exhibit increased fecundity and egg viability in warmer temperatures. *T. Cruzi* as well may exhibit increased viability at warmer temperatures, suggesting that incidence of Chagas disease will likely increase with climate change. Medone et al. 2015 find that the changing climate will likely create more geographic areas that are suitable habitats for Chagas vectors correlates with the force of infection of acute Chagas disease in Argentina and Venezuela. They find that warmer temperatures are unfavorable to vectors; although current Chagas hotspots may see decreases with increased temperatures, the overall geographic distribution of vectors may shift as previously too-cold areas warm.

Since Chagas disease is primarily found in rural areas in Brazil, multi-decadal trends in the urban and rural population may be a mediator in the future trajectory and control of Chagas disease (Delazeri, Da Cunha, and Oliveira 2022; Perz 2000; Randell and VanWey 2014). Brazil has become increasingly urbanized since the 1960s; internal migrants from rural, Chagas-endemic areas have resulted in identification of both acute and chronic cases of Chagas disease in places where Chagas is not historically found (Coura and Borges-Pereira 2010; Martins-Melo et al. 2014; Moncayo and Silveira 2009). Like all urban areas in Brazil, larger municipalities in Chagas-endemic areas have been increasing in size faster than the nearby rural areas, many of which have even seen population declines. The identification and control strategies of the 1980s that targeted known areas of Chagas endemicity may not be as effective for identifying latently infected internal migrants who have moved to the larger cities ourside of endemic strategies. As well, declines in the rural population are fundamentally changing the spatial distribution of new cases of Chagas disease. Although the rural population is declining overall, declines are not uniform across all areas of Chagas endemicity, and projections of future cases must include population change as well.

To identify areas of persisting and future Chagas endemicity, we borrow Bayesian disease mapping methods for modeling higher-incident spatially-clustered diseases, such as cancer (Best, Richardson, and Thomson 2005; Napier et al. 2019; Riebler et al. 2016; Wakefield 2007; Wikle, Berliner, and Cressie, Noel 1998; Knorr-Held 2000; Knorr-Held and Best 2001) and adapt these methods to suit the highly rare nature of Chagas disease. Since the rates of ACD are low—on the order of hundreds of cases annually for a population of over 200 million—and present in a highly clustered pattern in certain regions of Brazil, a specialized modeling approach is needed to highlight the spatio-temporal structure of Chagas disease. We use a Knorr-Held (2000) spatio-temporal model adapted with a rare count, zero-inflated model (Lambert 1992; Lee et al. 2016; Rathbun and Fei 2006; Ver Hoef and Jansen 2007) and include climate covariates and population growth to analyze how patterns of Chagas disease might play out for the ensuing decade. We find that with an increasing population and climate trends, it is likely that cases of Chagas will continue to increase in the absence of additional intervention, potentially as high as doubling in incidence.

## 2 Methods

### 2.1 Data

Counts of Acute Chagas Disease (ACD), aggregated by municipality ^1^ of residence and year between 2001 and 2019, are collected by the Ministry of Health’s Departamento de Informática do Sistema Único de Saúde (DATASUS; Department of Informatics of the Unified Health System) and retrieved from the agency’s TABNET database (Ministério da Saúde, Brasil 2023). We chose to use municipalities—the finest level of geographic aggregation available—in order to analyze spatial variability that may be lost at larger levels of geographic aggregation, like state or region. Over the period 2001-2019, 5568 cases of Acute Chagas Disease were reported among residents of 826 municipalities, with the highest counts in the northern states^2^ of Pará and Amapá. The municipality-specific total counts of Acute Chagas Disease reports are displayed in figure 1. Official population estimates at the municipality level are taken from Brazil Instituto Brasileiro de Geografia e Estatística (IBGE)’s annual population estimates for 2001-2006, 2008-2009, and 2011-2019 and the 2010 census counts (Instituto Brasileiro de Geografia e Estatística 2023). No data are present for 2007; population for this year is taken as the linear interpolation between 2006 and 2008. Climate data are retrieved from the European Union’s Copernicus Climate Change Service (C3S) Climate Data Store (CDS) Global Bioclimatic Indicators from 1950-2100 Derived from Climate Projections (Wouters et al. 2021), which contains a suite of 19 bioclimatic variables averaged annually. These variables are the same as those in the WorldClim (Fick and Hijmans 2017) dataset^3^, and are listed in supplementary table S2; a selection are displayed in figures 2 and 3. Broadly, we see in figure 2 that most reports of ACD occur in areas that are warmer (annual mean temp *>* 25^*°*^*C*) and wetter (annual precipitation above 2000 mm), although many cases do occur in drier climates. In Pará and Amapá, the UFs where most cases of ACD occur, there is a slight trend towards warmer and wetter weather, although substantial year-to-year variations present (figure 3). Projections are performed using the GFDL-ESM2M (NOAA, USA) algorithm.

**Figure 1.**
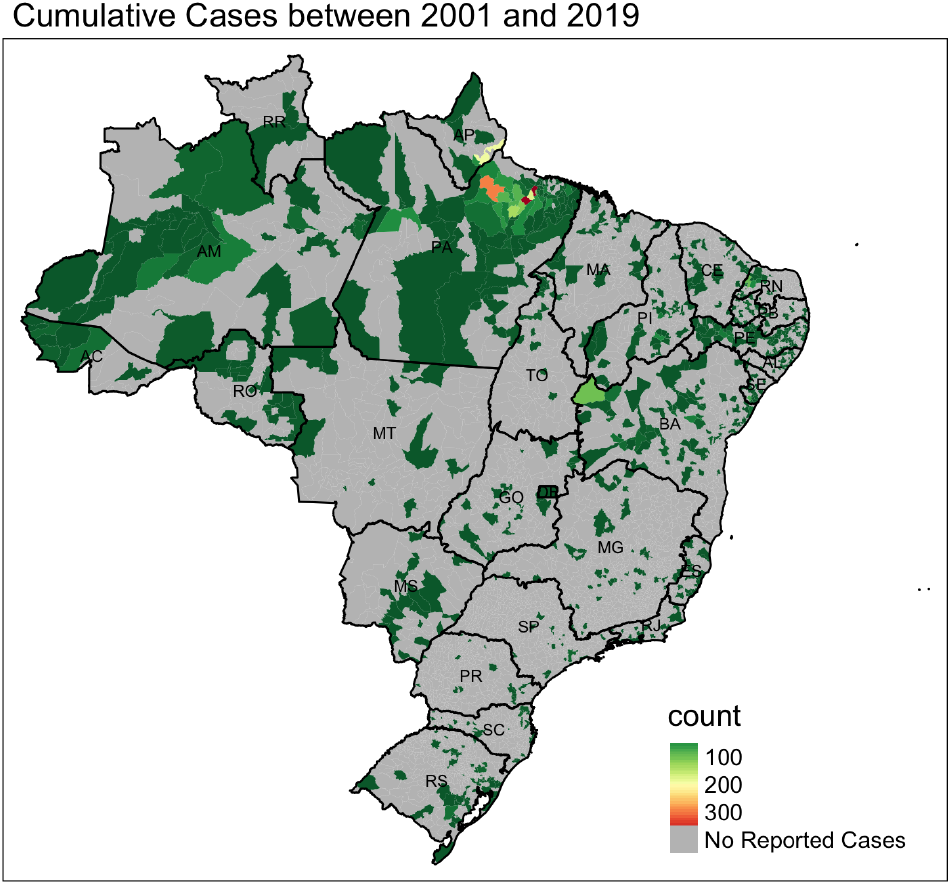
Counts of Chagas disease between 2001 and 2019 at the municipality level. Of the 5568 municipalities in Brazil, 4472 municipalities reported no cases of ACD during this period.

**Figure 2.**
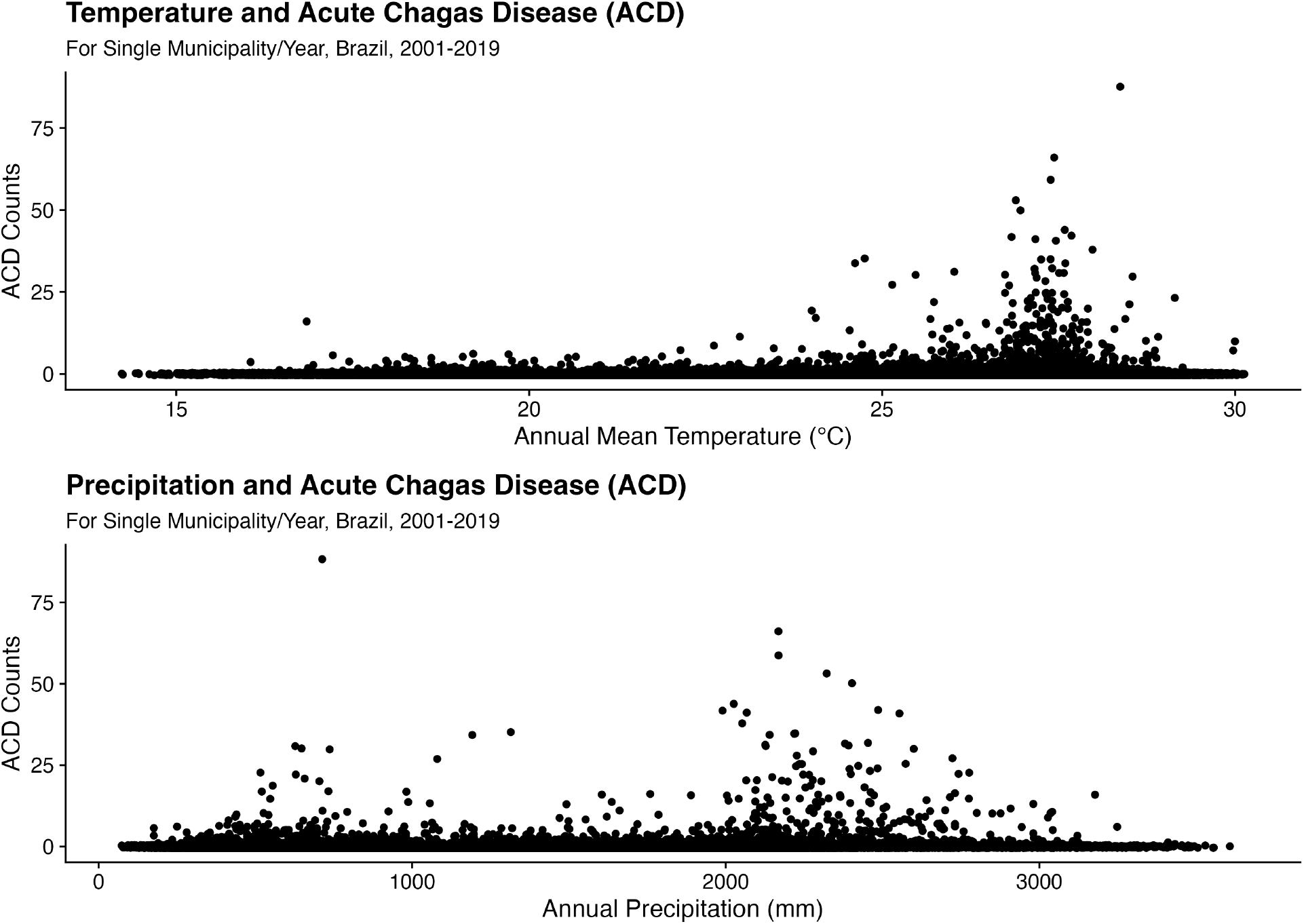
Counts of Acute Chagas Disease (ACD) by mean annual temperature and total precipitation, two of the 19 bioclimatic variables used in the analysis, displayed over the period 2001-2019.

**Figure 3.**
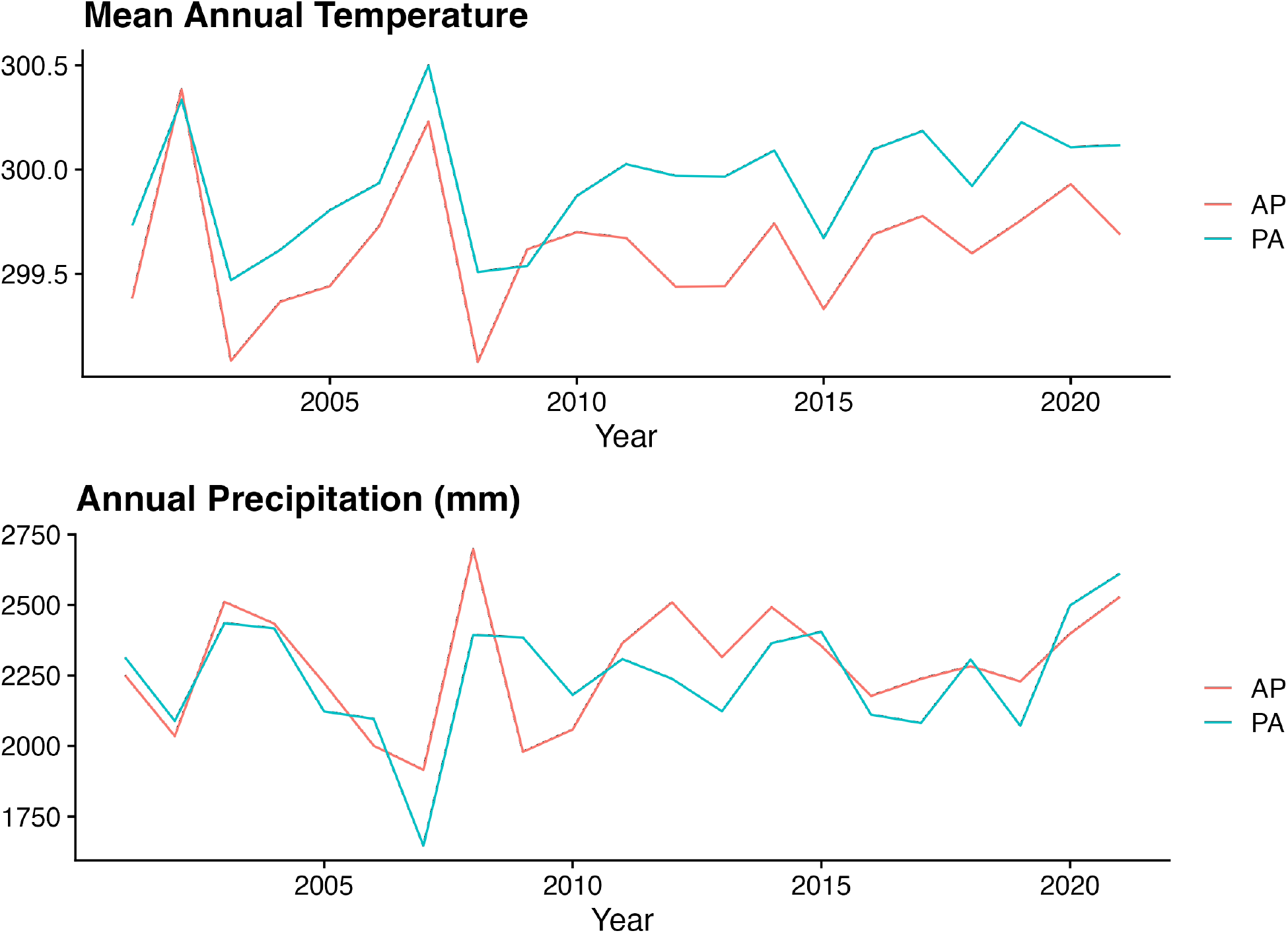
Mean annual temperature and total precipitation, two of the 19 bioclimatic variables used in the analysis, displayed over the period 2001-2030 for the two states with the highest incidence of ACD, Pará (PA) and Amapá (AP).

### 2.2 Conditionally AutoRegressive (CAR) statistical models for disease incidence data

Many classes of geospatial models for areal data (polygons, like municipíos) exist, a few of which are discussed here. Distributional models used in Bayesian modeling can be divided into two groups: Conditionally AutoRegressive (CAR) models, that describe probability for observations as conditional on their neighbors, and Simultaneous AutoRegressive (SAR) models, that use a matrix of adjacency weights to adjust for spatial dependence. At a high level, these two models differ in the scale of spatial dependence: the CAR model involves local smoothing, where the SAR model involves global smoothing. Here, we use a CAR model in our application to rates of Chagas disease, which is a highly local process found only in certain relatively isolated regions of Brazil and where global correlative structure is likely to over-smooth small-area variation. More commentary is provided in the supplementary material. As well, CAR models have a computational advantage over SAR models: they do not require matrix inversion, which can be computationally expensive or impossible when modeling thousands of small-area samples^4^.

The simplest implementation of the CAR model is the Intrinsically AutoRegressive (IAR or ICAR) model, also called the BYM model after authors Besag, York, and Mollie (1991). For a general Gaussian spatial process ***ϕ***, the CAR model is conditionally specified for each geographical unit as a normal distribution with expectation equal to the average of its neighbors and variance τ:

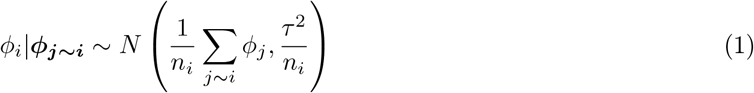

Where *ϕ*_*i*_ is an observation at the *i*th spatial unit, ***ϕ***_***j∼i***_ indicates the set of observations among the neighbors *j* of *i*, and *n*_*i*_ is the number of neighbors of *i*. Throughout, we refer to equation 1 as the CAR and IAR models interchangably. The IAR distribution can also be extended to Poisson, Binomial, and Logistic distributions as well (Besag, York, and Mollie, 1991; Haining, 2004).

To utilize the CAR distribution in a disease modeling context, Banerjee, Carlin, and Gelfand (2015) recommend using a pair of *random effects* for the standardized incidence rates of disease:

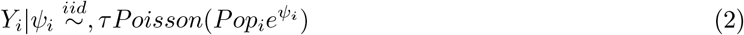

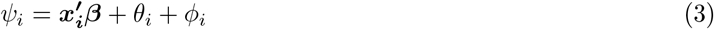

where ***x*** and *β* are vectors of spatially-varying covariates, *θ*_*i*_ captures heterogeneity with an *iid* normal prior *N* (0, 1*/*τ_*h*_) and *ϕ*_*i*_ captures spatial clustering with prior *CAR*(τ_*c*_) as in equation 1. Here, parameters *τ*_*h*_ and *τ*_*c*_ represent precision. Dividing extra-Poisson variability into ‘heterogeneity’ and ‘clustering’ poses a problem: should *τ*_*h*_ and *τ*_*c*_ be too large, the model will be unable to identify the two random effects. Indeed, priors on variance must be carefully chosen in order to allow for identifiability of *θ* and *ϕ*, which poses an existential question as to the utility of these models in the first place. Instead, other specifications including Leroux, Lei, and Breslow 2000 and the very closely related BYM2 model (Riebler et al. 2016) as implemented by Morris et al. 2019, which introduce a convolution of the spatial and aspatial error terms which allows for identifiability. The BYM2 model is a similar Poisson-GLM framework to the original BYM model, but replacing the pair of random effects with a convolved error term:

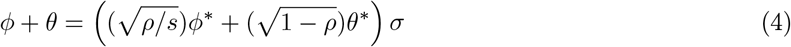

where *ρ∈* [0, 1] represents the proportion of variance that comes from the spatial clustering random effect and how much comes from the heterogeneity random effect; *ϕ*^*\**^ is the ICAR distribution; *θ*^*\**^ *∼ N* (0, *n*) where n is the number of connected subgraphs (in our application *n* = 1), s is the scaling factor such that *V ar*(*ϕ*) *≈* 1 (Riebler et al. 2016); and *σ >* 0 is the overall standard deviation for the combined error terms (Morris et al. 2019^5^). The BYM2 model improves upon the original form by allowing for independent definitions of the two prior distributions without involving *ρ* in the sampling process as is done for the proper CAR model. In doing so, this model involves the identification of only a single set of random effects rather than a pair of independent random effects, which improves identification by separating the dependency structure. Further, this avoids the need for informative priors in the BYM model as emphasized by Banerjee, Carlin, and Gelfand 2015. As well, in this context *ρ* has an informative interpretation, although it still does not map onto other indicators like Moran’s I. Morris et al. report that Stan’s Hamiltonian Monte Carlo (HMC) and No U-Turn Sampler (NUTS) provide faster and more precise inference with the BYM2 model than other samplers like WINBUGS and JAGS. The related Leroux (2000) model, which is similar to the BYM2 model but specifies the neighborhood matrix differently, has been shown through simulation to be superior to the original BYM model, and is employed by many in disease mapping studies (Lee, 2011).

#### 2.2.1 Extending the BYM model to include temporal effects

We follow the Knorr-Held (Knorr-Held 2000) framework for Bayesian spatio-temporal modeling. The Knorr-Held model adds time structure in a way that mirrors the BYM model (3) by adding temporally autoregressive effects *α*, temporal random effects *γ*, and a spatio-temporal interaction term *δ*:

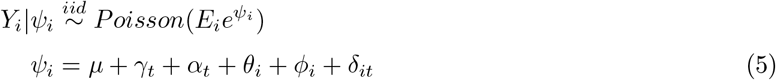

Where *μ* is the overall intercept, *γ* is an unstructured temporal component distributed *N* (0, *σ*_*γ*_), *α* is a structured temporal component that can be specified as an AR(1) or AR(2) process, and *δ* is a spatiotemporal interaction term. *ϕ* and *θ* are as described above. Effectively, the Knorr-Held model decomposes the overall pattern into a global temporal trend, a global spatial trend, and an interaction term between the two, in a procedure similar to ANOVA. Prior choice of *δ* is not straightforward, and requires careful thought about the relationship of space and time in the model. Knorr-Held (2000) lays out four types of priors, depending on the hypothesized interaction of the spatial and temporal dimensions: (I) where all interaction terms are *a priori* independent; (II) where interactions are autoregressive in time but independent in space;(III) where interactions are autoregressive in space but independent in time; and (IV) where interactions are totally dependent in both space and time. Further description of these interaction types is given in appendix section S4. Knorr-Held’s (2000) evaluation includes specification of the same disease model with each of the four interaction types, and evaluation of the resultant model by DIC.

#### 2.2.2 Zero-Inflated Poisson models for rare counts

Since Chagas disease is very rare, most entries in our matrix of counts by municipality and year are zero. While a low-rate Poisson may be able to capture this overdispersion of zeros, a more appropriate specification involves the zero-inflated Poisson model (ZIP; Lambert 1992). The zero-inflated Poisson is a mixture model that includes a Bernoulli process generating zeros and a Poisson process that generates counts (but may also generate some zeros). In this way, the zero counts in the data are effectively split into ‘structural’ zeros, which are generated from the presence or absence of the process of interest, and ‘sampling’ zeros, which are true random zero-counts in the presence of the Poisson process. Lambert (1992) note that in simulation, Poisson-only models are sufficient for a dataset that contains at most 68% zeros and 3.4% counts greater than 9, and that the ZIP model may be justified on datasets with higher rates of zeros. In our application to Chagas disease, over 99% of municipality-years have a zero count; nonzero entries have an average of 1.65 (95% CI: 1-7) infections. The ZIP distribution is parameterized by Bernoulli probability π and Poisson rate *λ*:

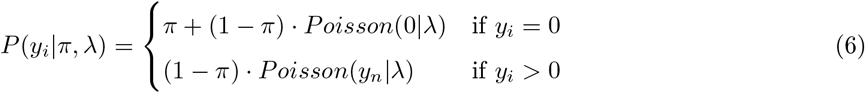

The ZIP distribution is appropriate in a GLM framework, where parameters *π* and *λ* are specific to each observation *y*_*i*_ and are estimated with logit and log link functions, respectively. When writing the probability statement, we can also take advantage of the fact that *Poisson*(0|*λ*) = *λ*^0^*e*^*−λ*^*/*0! simplifies to *e*^*−λ*^, clarifying the condition where *y* = 0:

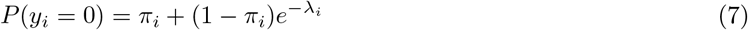

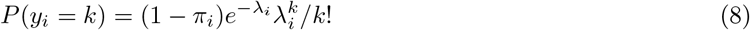

In turn, the central moments of the ZIP distribution are mean (1 *− π*)*λ* and variance *λ*(1 *− π*)(1 + *πλ*) (Lambert, 1992).

The ZIP distribution then prompts an additional modeling decision. When used in a GLM framework, covariates can be added to both the Poisson process and the Bernoulli process. Prior studies have done both: Agarwal, Gelfand, and Citron-Pousty 2002 include a spatially-autocorrelated Poisson process, Rathbun and Fei 2006 include a spatially-dependent Bernoulli process, and Ver Hoef and Jansen 2007 include spatial (and temporal) autocorrelation in both parts. For our application to Chagas disease, it is not clear if the spatial process should be included in either the Bernoulli or Poisson process, or in both parts. Is the probability of any appearance of Chagas disease spatially correlated? Undoubtedly, as the primary determinant in risk of Chagas disease is the highly localized distribution of *T. Cruzi*. However, conditional on the presence of *T. Cruzi*, it is less clear *a priori* if the risk of contracting Chagas also a spatially dependent process.

### 2.3 Model: Estimating rates of ACD with zero-inflation and spatial and temporal autoregression

Here, we integrate the components discussed above into a single Bayesian model that allows for spatial and temporal autoregression as well as overdispersion of zeros. Specifically, we use a ZIP likelihood within a spatio-temporal decomposition framework like the one proposed by Knorr-Held (2000). As well, we innovate by introducing the BYM2-type convolution of the unstructured error and spatially-structured heterogeneity to improve identification on the model posed by Knorr-Held (2000). We encountered convergence issues when including the spatial convolution in the Poisson proccess; as a result; this process is defined in the Bernoulli parameters only. In the Poisson process, we include a grand mean, temporally AR(1) time trend, and a spatial fixed effect with a Knorr-Held Type 1 interaction. We chose the probabilistic programming language and software suite `Stan` to estimate the yearly incidence risk of Chagas disease across all municipalities in Brazil between 2000 and 2019. The model is evaluated in `Stan` 2.20 using the `cmdstanr` (Stan Development Team 2023) interface for the `R` programming language, version 4.20. `Stan` was chosen for its speed relative to other probabilistic programming languages, like GeoBUGS (Lunn, Arnold, and Spiegelhalter 2004) or JAGS (Plummer 2003), especially for its ability to evaluate vectorized probability statements. Although Stan lacks the built-in support for spatial models present in BUGS, the computational gains from vectorization and adaptive sampling allow for quick evaluation and convergence of complicated posteriors, with full implementation details elaborated in the supplementary material. To sample Bayesian posteriors, by default Stan uses the No U-Turn Sampler (NUTS), a variant of Hamiltonian Monte Carlo, in contrast to Gibbs sampling used by BUGS and JAGS.

We run two formulations of the model: first, a non-covariate smoothing model used purely to recover latent rates of Chagas disease unadjusted for other causal factors besides population at risk; and second, a model that includes climatological covariates. The two models differ only in the inclusion of the set of covariates.

Beginning with ZIP-distributed likelihood:

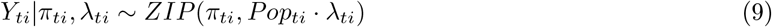

Where indices *t* and *i* refer to year *t* between 2000 and 2018 and municipality *i* between 1 and 5561, the number of municipalities in Brazil. Assuming a logit link for Bernoulli parameter *π* and Poisson parameter *λ*, we take the follow GLM equations for *π* and *λ*:

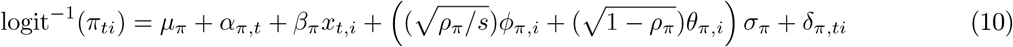

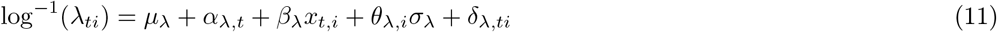

Where *μ* indicates the global mean with uninformative, *U* (*−*20, 20) prior; *α* is an AR(1) structured time effect; *ϕ* is a structured spatial process with an *IAR*(1) prior; *θ* is an unstructured spatial error with an independent *N* (0, 1) prior; *ρ* indicates the proportion of variance that comes from the spatially structured process, with prior *Beta*(1*/*2, 1*/*2) (Mitzi Morris 2018); *σ* is the variance of the convolved spatial term; and *δ* is a spatio-temporal interaction with a normal prior at mean 0. Finally, *β*.*x*_*t,i*_ indicates a set of coefficients and covariates, which are absent in the main smoothing model but include a set of environmental covariates in the climate model. For simplicity, the effect of these covariates is assumed to be constant throughout space and time. We follow Knorr-Held’s recommendation to drop the unstructured temporal component *γ* to improve identification of the model and the parameters in equation 10 are otherwise same as described in equation 5. Finally, we specify uninformative Gamma-distributed hyperpriors for variance as recommended by Knorr-Held 2000:

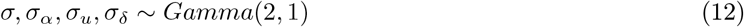

The quantities of interest include the expected number of acute Chagas Disease cases in year *t* in municipality *i*, which is given by:

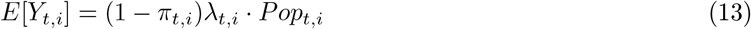

And the incidence rate:

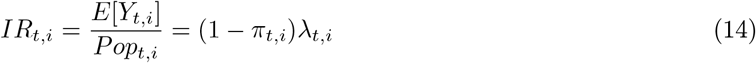

Altogether, this model was evaluated in Stan using the CmdStanR interface for the R-language. Four chains were in parallel run for 2000 warmup iterations and 1000 posterior draws, and evaluated in approximately 12 hours on an Apple MacbookAir M1. The corresponding Stan code is included in the supplementary material.

#### 2.3.1 Climate Model

To investigate the relationship of climate with Chagas incidence, we include covariates to the model related to temperature, precipitation, and vegetation. Determining covariates relevant to the incidence of Chagas disease is not straightforward as the climate processes that affect *Triatomines* and the *T. Cruzi* parasite may not necessarily be the same as those governing transmission to humans. Further, interventional strategies limiting transmission have shown an overall decrease in incidence of Chagas disease, which may confound identification of climatic factors influencing transmission. Nonetheless, we have chosen to include the following covariates in our model.

In laboratory settings, it was found that *Triatomines* incubated at warmer temperatures (30C vs. 28C and 26C) mature faster and had higher levels of *T. Cruzi* parasites in stool, although insect mortality did increase slightly (Tamayo et al. 2018). Further, *Triatomines* may be able to adapt to changes in temperature in complex ways (Clavijo-Baquet et al. 2021). Ecological modeling of Chagas Disease in North America indicates that as temperatures rise, the distribution of *Triatomines* may shift towards the north and northeastern part of the region (Garza et al. 2014).

To avoid multi-collinearity among our climatological factors, like Medone et al. 2015 we use Principal Component Analysis (PCA) on the 19 WorldClim Bioclimatic Indicators (Fick and Hijmans 2017), which we retrieved from the Copernicus Climate Change Service (C3S) Climate Data Store (CDS) dataset, “Global Bioclimatic Indicators from 1950-2100 Derived from Climate Projections” (Wouters et al. 2021). PCA is a dimensionality-reducing procedure that decomposes the matrix of covariates by municipality-year into an ordered set of orthogonal vectors, or principal components. Each principal component represents a ‘trend’ or pattern in the data with the first component representing the most dominant pattern by proportion of variance explained, and each subsequent component representing less of the variance. Each observation can then be described as a linear combination of principal components and coefficients. PCA can be used for Principal Component Regression; rather than using the covariates directly, each observation’s location in principal component-space is used as a covariate. Since all principal components are orthogonal, this avoids any potential multi-collinearity in the data.

We find that the first six principal components explained 95% of the variation in the data. Values for these principal components are displayed in supplementary table S3 and the variance explained in supplementary figure S3. The largest principal component, responsible for just over 50% of the variance in the dataset, is related to warmer, drier weather year-round. In turn, the second principal component (17% of total variance) is related to cooler temperatures with more seasonal fluctuation but less precipitation year-round. Third and subsequent principal components are less clear in their interpretation and are responsible for decreasing amounts of variance in the dataset.

#### 2.3.2 Projection of future incidence

The resulting quantities estimated from the main smoothing model and the covariate model are used to estimate the incidence of Chagas disease over the 10 year period from 2020 to 2030 using projected population counts and projected climate variables. To project this data, we calculate the average annual exponential growth rate over the period 2001-2019 for each municipality as:

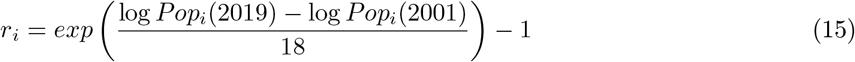

Growth rates at the municipality level are displayed in figure 4. The overall growth rate of Brazil is 1.04% per year between 2001 and 2019, and the average municipality grew by 0.6%. The 2019 population for each municipality is projected forward each year for 10 years as:

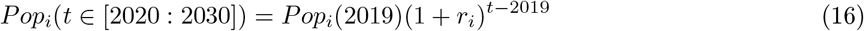

**Figure 4.**
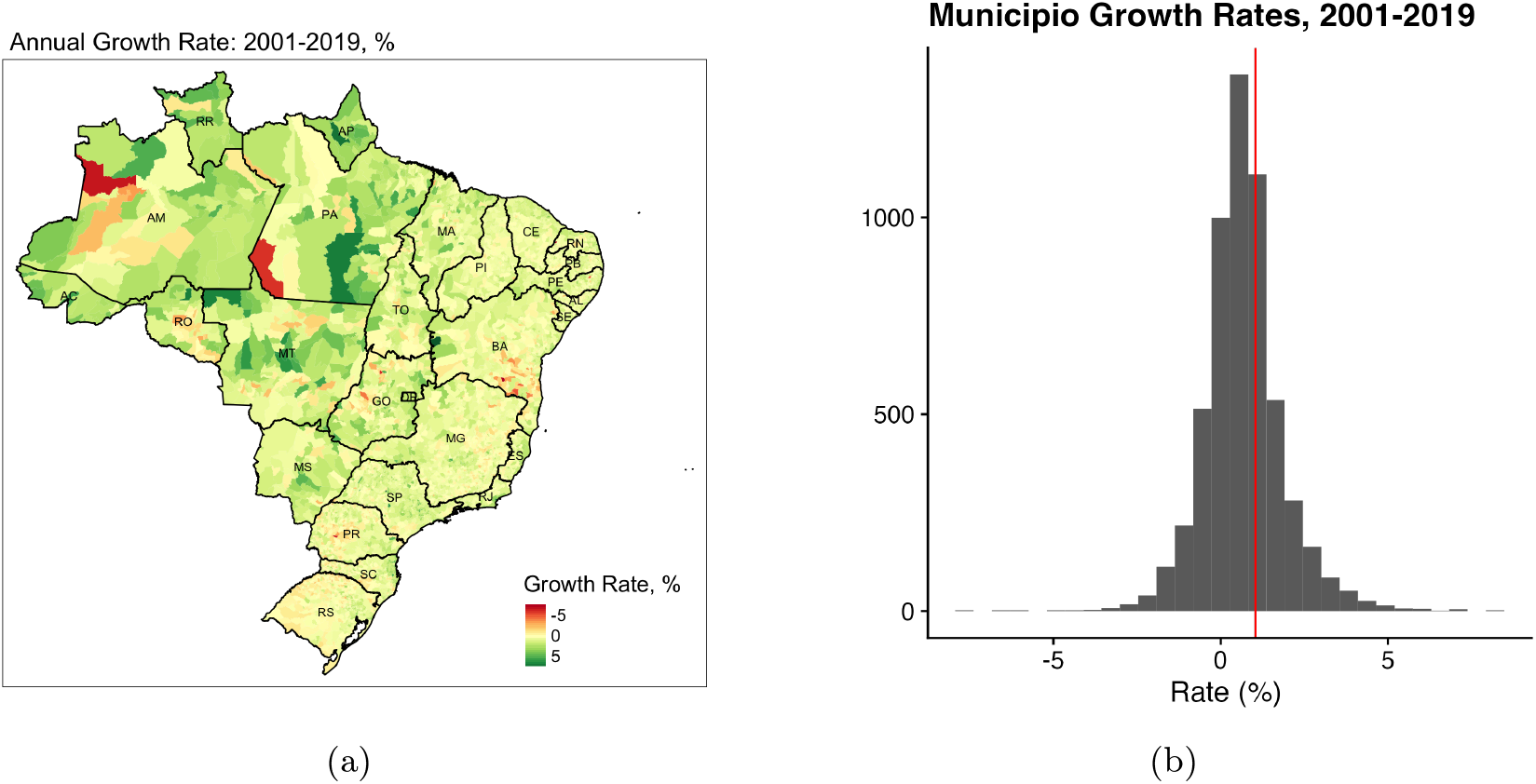
(A): Average annual percentage change in population for all municipalities in Brazil between 2001 and 2019. (B): Average annual growth rate, percent, for all municipalities in Brazil between 2001 and 2019. The overall growth rate for Brazil, indicated in red, is 1.04%, higher than the average municipality growth rate of 0.6%.

This projected population is used as an input to the model to predict future incidence of Chagas disease. We report two sets of projected rates of Chagas disease: one from the main smoothing model and one from the covariate model. In the covariate model, each municipality-year’s location in prinicipal component space for the predicted bioclimatic variables is used as an additional input. The incidence in municipality *i* at future year *t⋆* is estimated as:

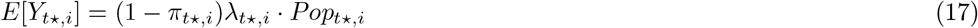

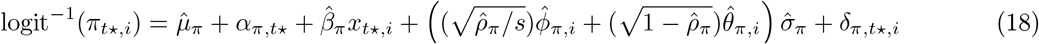

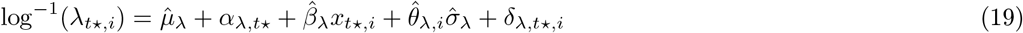

Where all quantities are the median value of those estimated in the main model except time-trend quantity α. for both α_*π*_ and α_*λ*_, which is taken as an AR(1) random walk given the distribution of estimated time trend terms:

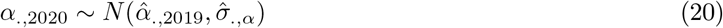

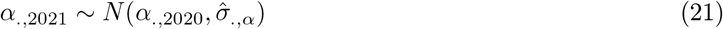

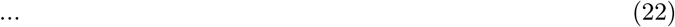

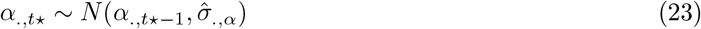

We conduct 1000 simulated random draws of future incidence and report the predicted rates of Chagas disease.

## 3 Results

### 3.1 Results of Main Smoothing Model

The overall incidence rate of Acute Chagas Disease in Brazil between 2001 and 2019 is estimated to be 0.121 per 100k person-years of life (PYL), although substantial heterogeneity in risk exists between and within regions. Figure 5 shows the municipality level 18-year incidence rate of acute Chagas Disease. Estimated incidence of Chagas disease is highly spatially variable with a strong regional trend, with two major areas of vulnerability: first, the northern Amazon states of Amapá (AP) and Pará (PA), which have the highest smoothed incidence rates in the country at 1.80 and 1.69 per 100k PYL—almost an order of magnitude higher than the national average—as well as Acre (AC; 0.317 per 100kPYL) and Amazonas (AM; 0.188 per 100kPYL). These states are highly rural and have a smaller population than the coastal states, but contain the majority of Acute Chagas Disease risk. The second main region of transmission includes the northeastern, Caatinga states of Rio Grande do Norte (RN; 0.334 per 100kPYL), Sergipe (SE; 0.247 per 100kPYL), Piauí (PI; 0.197 per 100kPYL) and Pernambuco (PE; 0.316 per 100kPYL). We do not observe increase transmission rates in the Cerrado, which includes the state of Goiânia and Mato Grosso do Sul, as reported by Gurgel-Gonçalves et al. 2012 besides a slight elevation in Tocantins. The states with the lowest estimated rates of Chagas Disease are the federal district of Brasilia and Sao Paulo.

**Figure 5.**
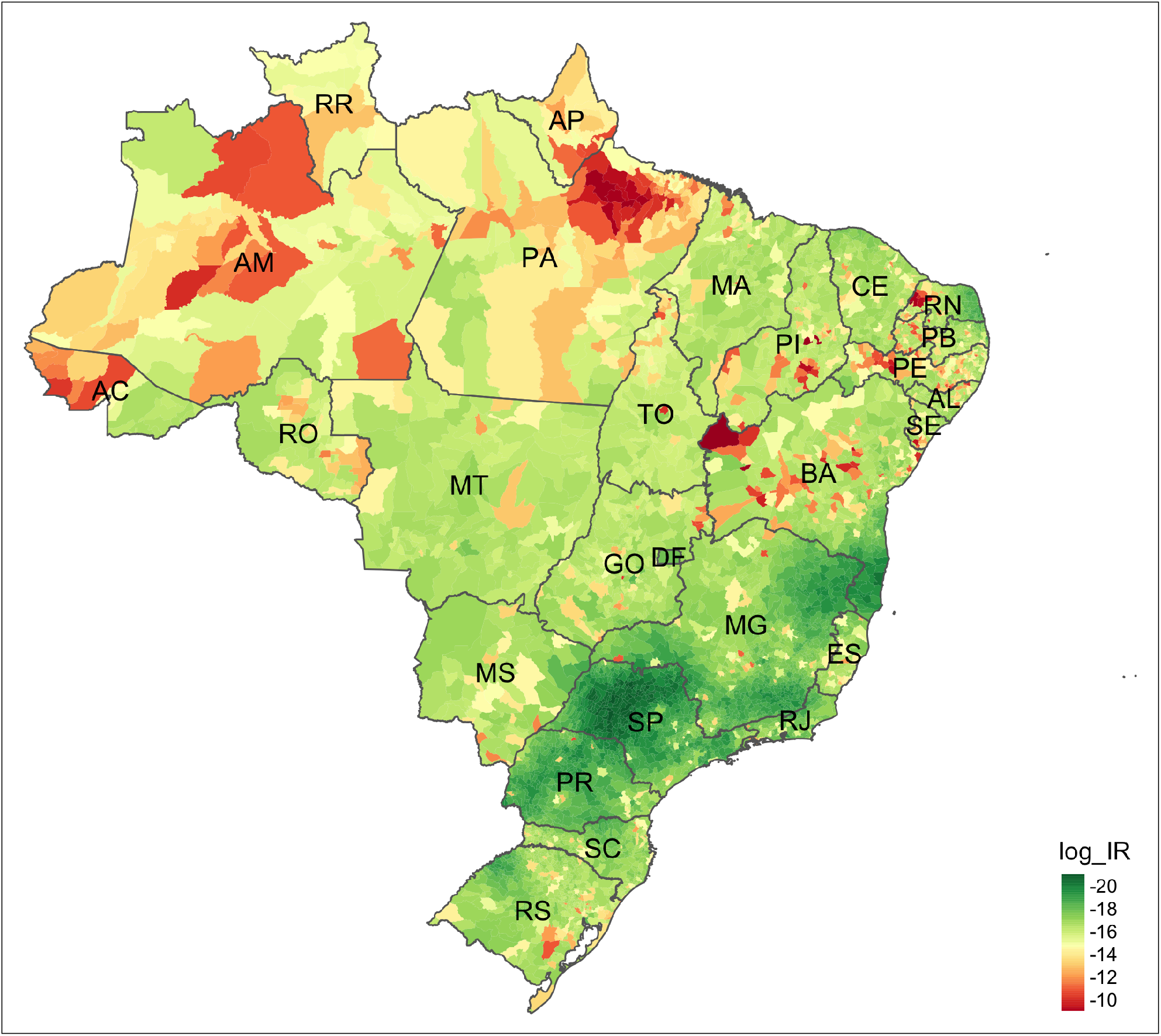
Overall log incidence rate at the municipality level over 2001-2019. Red indicates higher rates of ACD and green indicates lower rates of ACD. Incidence rate is calculated as log 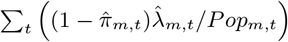 .

Within the Amazon states of Pará and Amapá, which have the highest overall rates of new Acute Chagas Disease diagnoses, 31 of 160 municipalities had 18-year incidence rates higher than 1 per 100k PYL; the highest rates of ACD were found in Breves (population 93,000) and Limoeiro do Ajuru (25,000), with 15.9 and 15.8 cases per 100k PYL respectively. Six more municipalities had rates above 10 per 100kPYL: Curralinho, Abaetetuba, Bagre, Muaná, Anajás, and São Sebastião Da Boa Vista. However, the most cases were predicted to be found in Belém, the capital and largest city in Pará, at 386 over the 18 year period for a population of approximately 1.4 million.

A zero-inflated models represent the observed data as being a mixture of two processes: here, the probability of never being exposed to Chagas disease represented through the Bernoulli process, and the incidence rate given exposure represented through the Poisson process. Lambert 1992 refers to the over-dispersion of zeros generated through these processes as these as ‘structural’ and ‘non-structural’/’sampling’ zeros, respectively. Although we found a very strong spatial process governing the rate of ‘structural’ zeros—probability of never being exposed to Chagas disease (shown in figure 6)—we did not find a strong spatial process in the rate given exposure. Since Chagas disease is transmitted to humans given contact with disease-transmitting vectors with a particular habitat, we interpret this to mean that Chagas-carrying *Triatomines* are more likely to live in certain locales, the rate of contact and transmission within those locales is more spatially constant.

**Figure 6.**
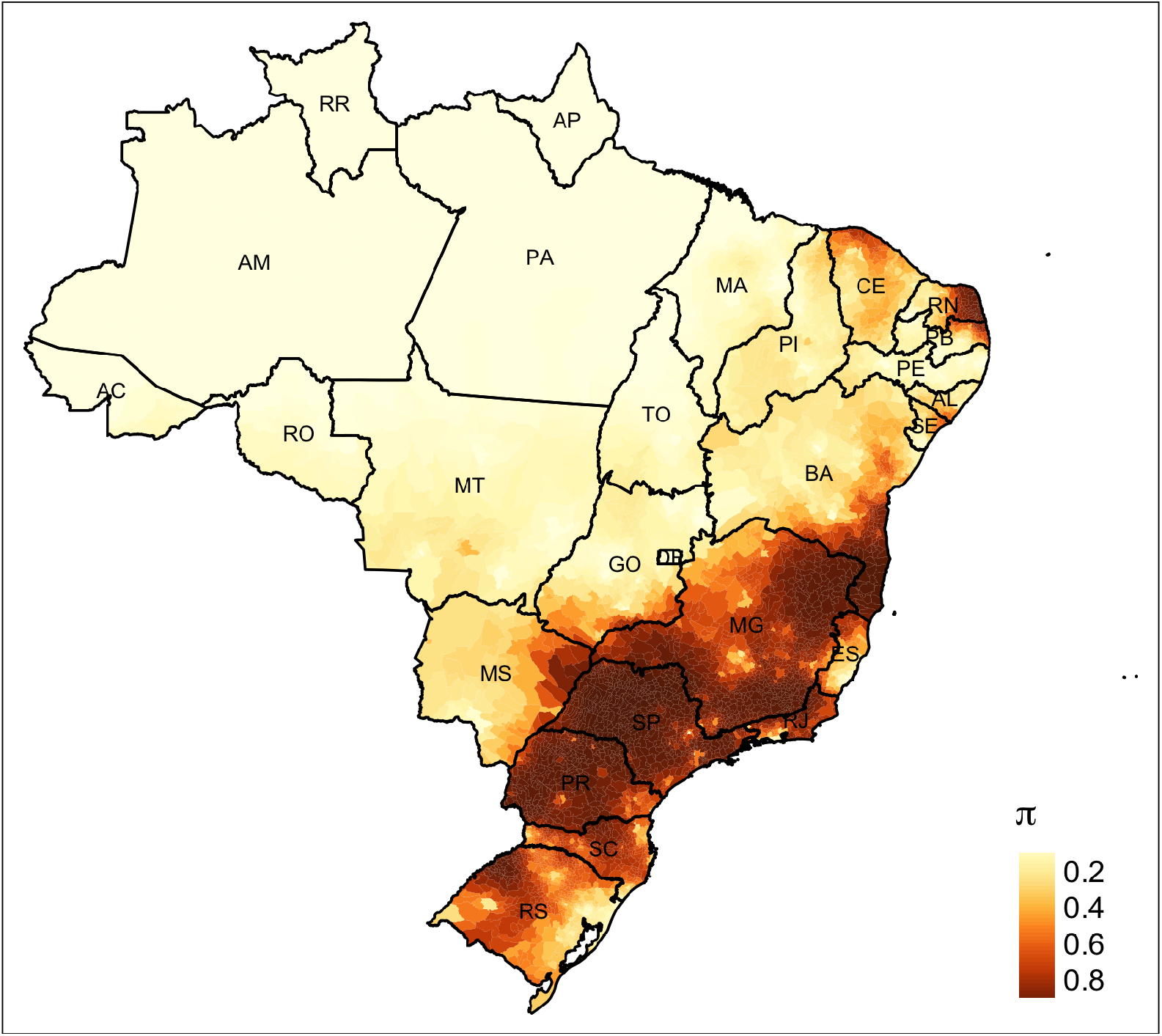
Estimated spatial process *π* governing the probability of an individual never being exposed to Chagas Disease over the study period. This process is an inverse-logit transformation of a linear combination of Conditionally AutoRegressive (CAR) term *ϕ*_*π*_ capturing risk that is spatially clustered, possibly due to Triatomine habitat or contact rates.

Overall, the Northern and Amazon states were found of have a high probability of exposure to Chagas Disease and the coastal and southern states were less likely to have exposure. The total spatial term for *π* is shown in figure 6. Parameter *ρ*, indicating the proportion of spatial variance derived from the ICAR term, evaluated to 0.985, indicating that the spatially-clustered process was responsible for most of the Chagas incidence and the random process *θ* contributed very little to the overall distribution, indicating further that the location of *Triatomines* may be driving the location of cases.

Where the probability of exposure to Chagas disease shows a strong spatial pattern, the rate of Chagas— after normalization for population—does not show nearly the degree of spatial autocorrelation as the Bernoulli process. The spatial structure for the Poisson process *λ* that estimates the rate of disease is shown in supplementary figure S1. Instead, even though the Poisson process parameter *λ* is normalized to municipal population, the Poisson process instead appears to be highlighing population locations rather than a spatially-autocorrelated process. However, Moran’s I test for spatial autocorrelation did find that the spatial heterogeneity term *θ* was statistically significantly spatially autocorrelated, albeit weakly (*E*[*I*_0_] = *−*0.0001; *I*_*a*_ = 0.12; *p*(*I*_0_ *< I*_*a*_) *<* 2*e −* 16). Future evaluations of this model will need to carefully consider how to incorporate autocorrelation into the Poisson process while maintaining model identification. The overall time trend parameters α_*π*_ and α_*λ*_, which are specified as AR(1) processes, both show a difference from 0 on the linear scale, indicating that there is a global temporal component in both the Bernoulli and Poisson processes (figure S2A). However, after adding in mean terms *μ* and applying the logit and log transforms as shown in figure S2B, the overall time trend tells a different story: the Bernoulli probability *π*, indicating probability of non-exposure to acute Chagas Disease, decreases from 55% in 2000 to a maximum of 23.6% by 2006 only to increase to nearly 100% for the remainder of the study period. This indicates that over the course of the study period, country-wide exposure to Chagas disease increased before decreasing to nearly 0 after 2007, at which point the distriubtion of Chagas cases ceased to be a country-wide phenomenon and instead became more spatially localized. The Poisson rate is stable around 2e-5 per capita over the course of the study period.

Overall, the model converged well and showed good mixing between the chains for the main parameters

*π*and *λ*. The Root Mean Squared Error (RMSE) of the model, evaluated as:

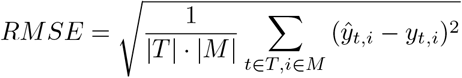

is 0.175. Convergence is evaluated using statistic 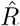, which evaluates the agreement of between-chain estimates, and Effective Sample Size (ESS), which evaluates the number of samples correcting for autocorrelation. Supplementary table S1 shows the distribution of 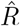 and ESS for all parameters *π, λ, ϕ, θ*, α, and *δ*.

### 3.2 Results of Climate Model

The climate model includes the specification as the main model above with the inclusion of each municipality-year’s location in principal component space among the 19 WorldClim Bioclimatic variables. We include the first six principal component dimensions as covariates *β* in both the Bernoulli process governing overdispersion of zeros and the Poisson process governing rate of Chagas disease. The RMSE of the climate model is 0.183, which is slightly higher than that of the main model, indicating that controlling for climate produces a worse fit and may reduce the accuracy of the model, possibly due to overfitting. Posterior densities of *β* are included in supplementary figure S4. In the Poisson process, posterior estimates of coefficients for the first three principal components were found to be statistically significantly different from zero, whereas in the Bernoulli process, only the second principal component was found to be significantly different from zero. The values of parameters in the climate model are similar to the values in the main model (shown in figure S5), except for *ϕ*_*π*_, the spatial clustering term in the Bernoulli process.

To better analyze these climatic factors, we transformed these estimated coefficients from principal component space back to the scale of the original variables before applying the inverse-link function and intercept terms to show the values visualized in figure 7. These values are calculated as 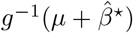, where *g* is the link function *log* for the Poisson process *λ* and *logit* for the Bernoulli process *π*, and 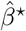 represents the estimated coefficients transformed from principal component space to the original coefficients. Ultimately, *β*_*π*_ coefficients represent a one-unit change in each variable on the probability of non-exposure to Chagas disease, and *β*_*λ*_ represents the effect of a one-unit change in each variable on the predicted rate of Chagas disease, per million person years, conditional on exposure. Overall, we see that these variables do not affect the rate of Chagas disease, only the probability of non-exposure. Non-exposure to Chagas disease is more likely in climates that are highly seasonal, and less likely in wetter wetter climates.

**Figure 7.**
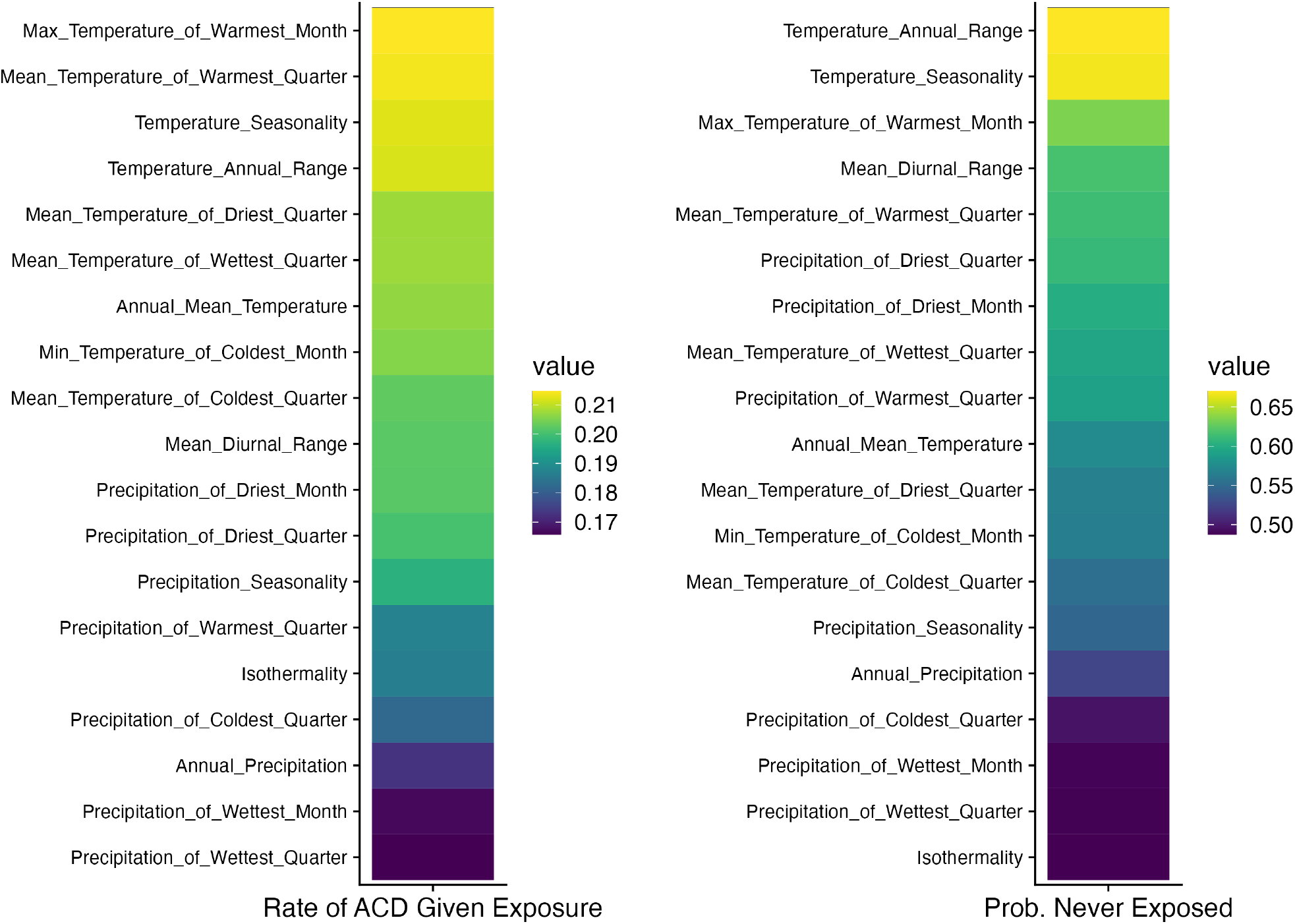
Coefficients for 19 WorldClim Bioclimatic Variables used in the climate model estimated for both the Poisson process (*λ*, left), governing the incidence rate of ACD given exposure, and the Bernoulli process (π, right), governing the probability of never being exposed to ACD. Lighter colors on the left figure indicate that a higher value of the coefficient corresponds to a higher rate of ACD in the population where exposure is present, and on the right figure indicate that probability of never being exposed is higher. Coefficients are estimated in principal component space and transformed to the natural scale, and applied the corresponding *log*^*−*1^ link function for the Poisson process and *logit*^*−*1^ for the Bernoulli process.

### 3.3 Projected Rates 2020-2030

We used the main smoothing and climate covariate models to estimate counts of Chagas disease over the decade 2020-2030. As elaborated above, the projection procedure utilizes the estimated intercept and spatial parameters, and using randomly-drawn temporal structure. The main model estimates a median of 4461 cases of Acute Chagas Disease over the decade 2020 (IQR: 1,653 - 13,859), almost double the number of cases in the previous decade (2,612). Predicted incidence is similar when including the bioclimate covariates, estimating a median of 4461 cases (IQR: 1619 - 13,270). Figure 8 shows the median annual predicted counts of Chagas disease across Brazil and interquartile range between 2020 and 2030. A map of projected incidence and a comparison of observed and projected rates are shown in figure 9 and for selected municipalities in figure 10 and 11. Most of ‘hot spots’ for new cases are predicted to be in the same locations as 2001-2019, including Abaetetuba, Belém, and Breves in Pará, and Macapá in Amapá. However, the largest increases are projected to be in smaller, rural municipalities with high growth rates in the states of Amazonas—especially municipalities Apuí and Tefé—and Piauí. The climate covariate model implies slightly lower rates than the main model, implying that projected bioclimatic conditions may result in fewer infections, although the overall difference is likely small. The trend observed in figure 8 shows a highly variable trajectory year to year—much more than the main model predicts—indicating that annual climate fluctuations may have substantial effects on predicted rates.

**Figure 8.**
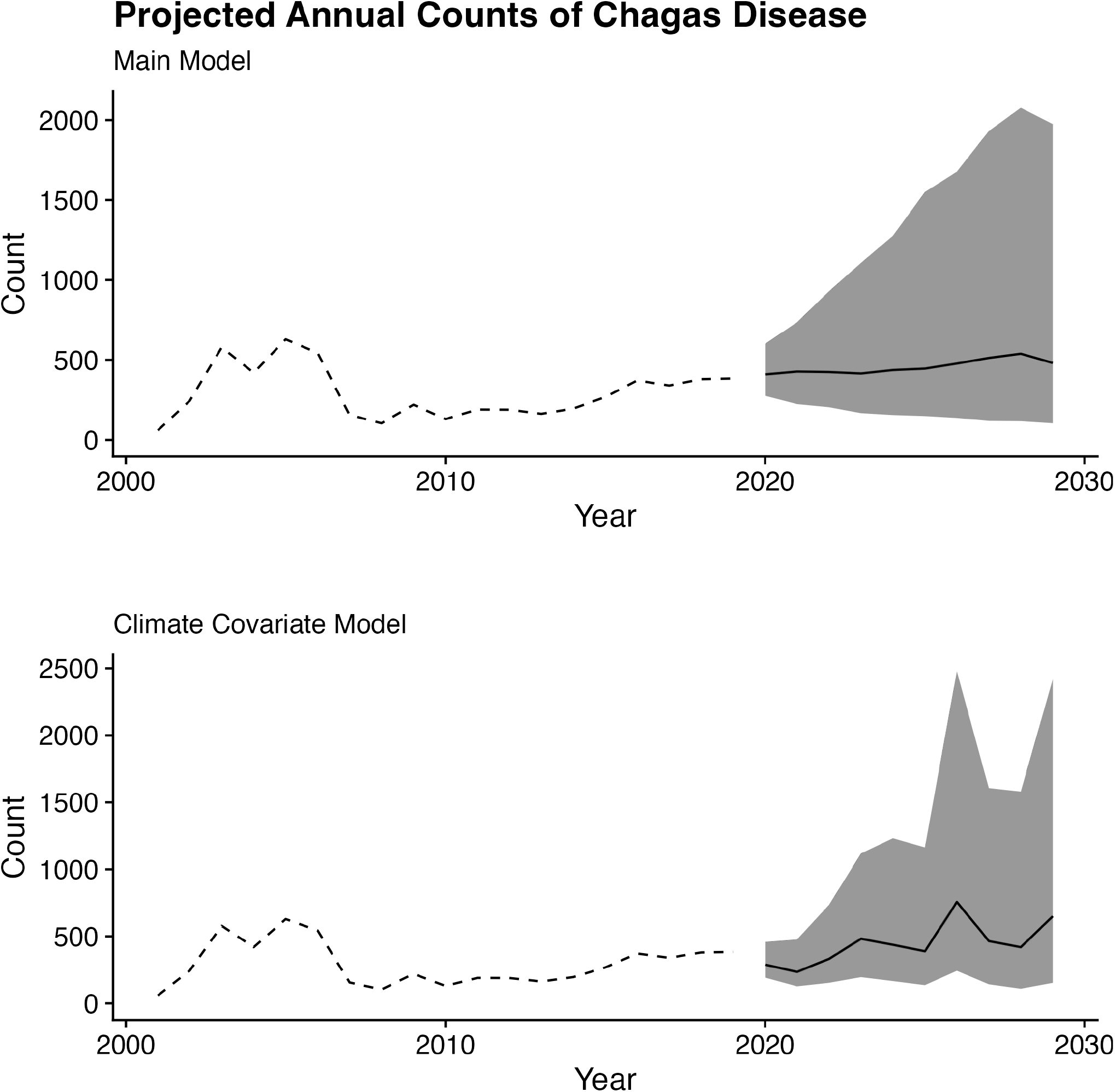
Observed counts across Brazil, 2001-2019, summary of 1000 projected counts, 2020-2030, from the main smoothing model (top) and climate covariate model (bottom). Median simulated counts are shown in black and interquartile range, representing 50% of simulations, is shown in grey.

**Figure 9.**
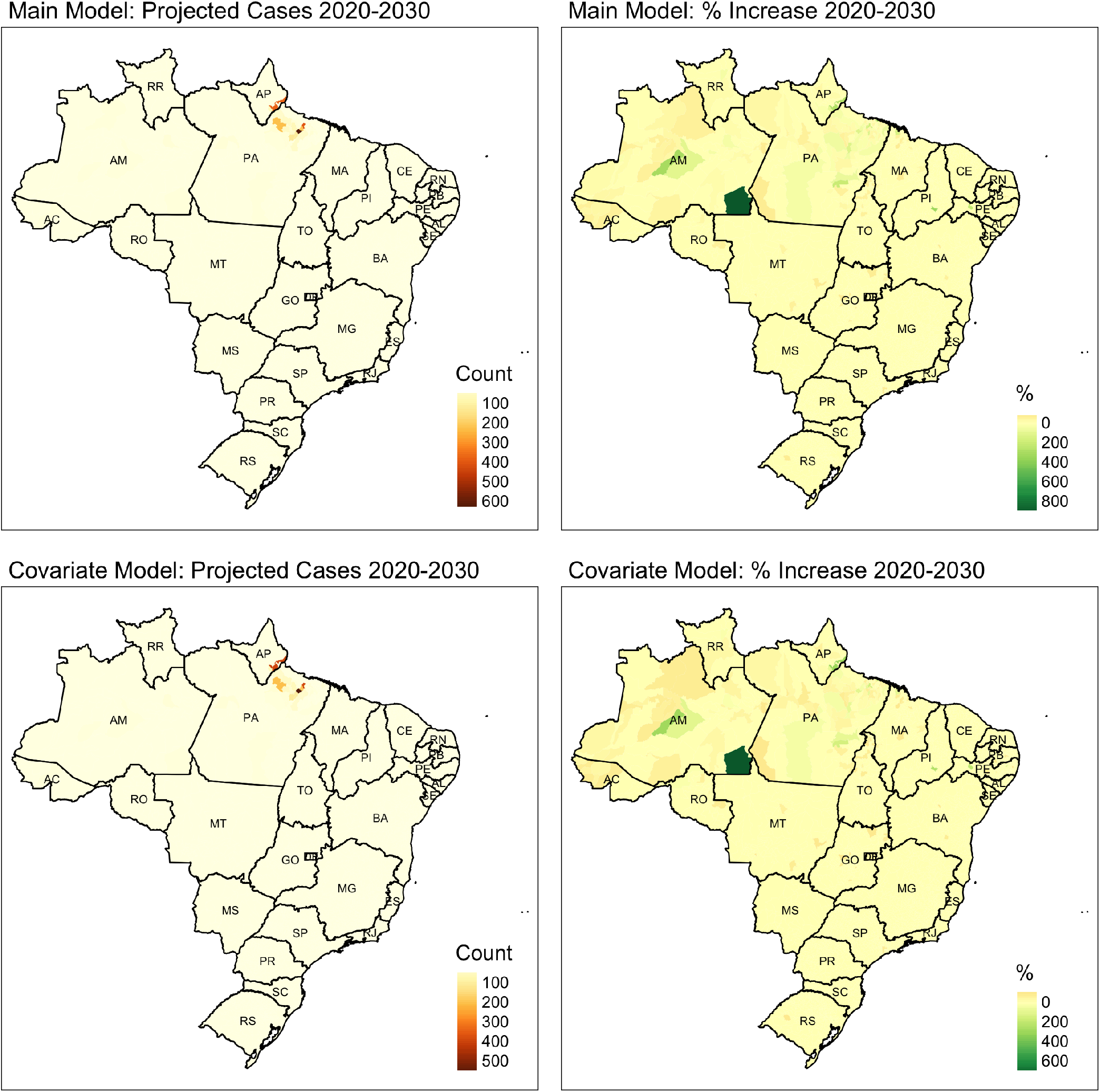
Projected incidence and percent increase compared to the previous decade.

**Figure 10.**
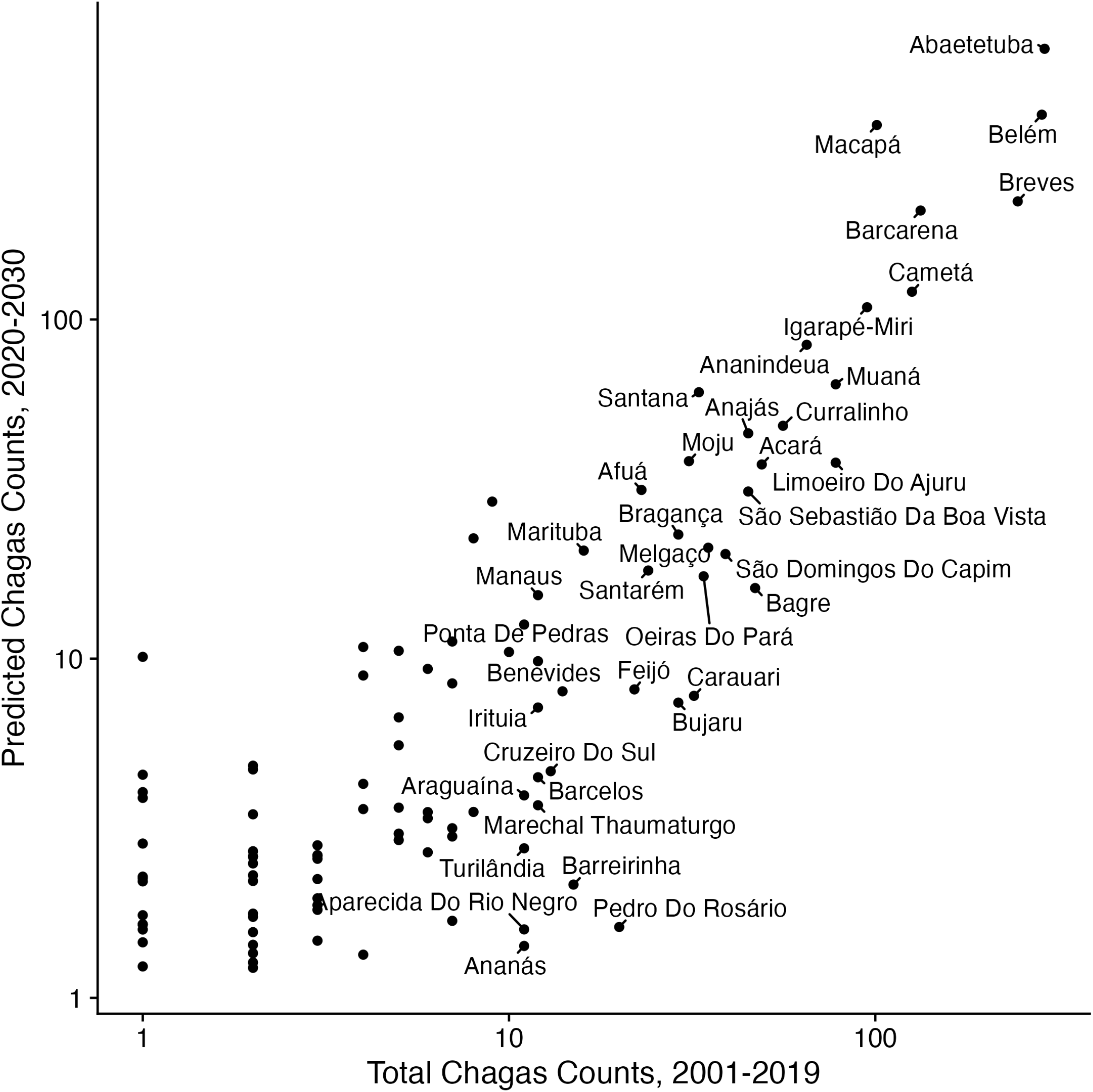
Observed and Projected rates, main model

**Figure 11.**
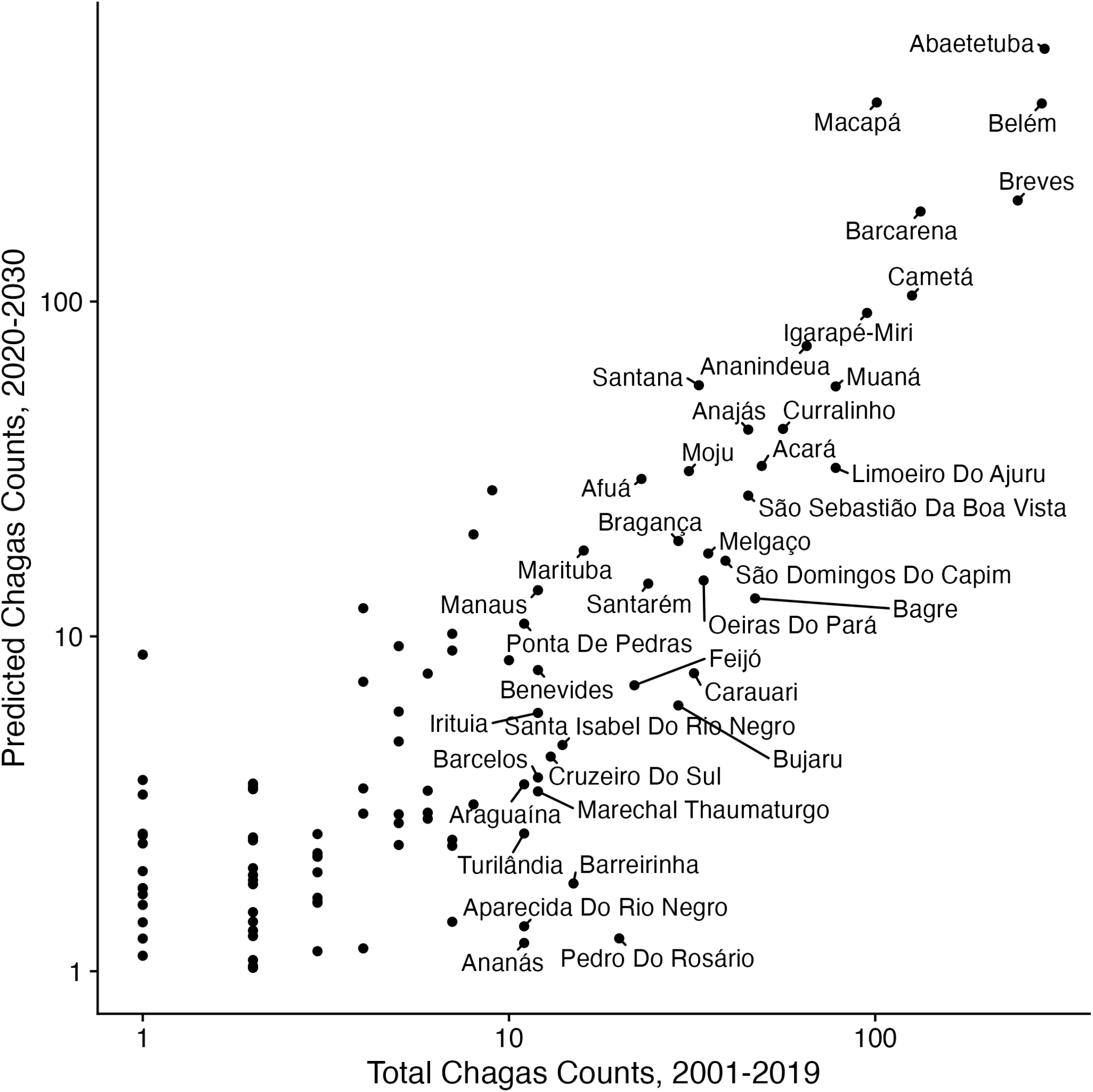
Observed and Projected rates, climate model

## 4 Discussion

Despite progress towards elimination, Chagas disease remains a significant threat to public health, and additional intervention will be needed to further reduce rates of new ACD cases in Brazil. The results of our modeling imply that rates of ACD will increase consistently over the ensuing decade—potentially as much as doubling compared to the previous decade—driven primarily by an increasing population in high-risk areas. Climate change may result in the exacerbation of this trend. However, this appears to be at odds with the country-wide decrease in ACD rates as a result of *Triatomine* eradication campaigns since the 1980’s. While these campaigns have been enormously successful, risk of ACD is likely to persist without additional intervention. As a result, our work here serves as a call to continue the campaigns Nonetheless, we predict that risk for ACD will persist, and further intervention will be necessary to continue the decrease observed in previous decades.

This model has a number of limitations, some of which may be addressable in future modeling studies. First, our data include official reports of Acute Chagas Disease as submitted to SINAN. We were unable to find literature estimates of under-reporting rates of ACD not submitted to SINAN. It is possible that there are many annual cases not captured in the dataset, implying that our estimates are not only incomplete, but subject to a ‘survivorship paradox’ where the locations of highest epidemiological interest are not captured in the dataset. Since the treatment success rate is high for acutely diagnosed cases of Chagas disease, it may be reasonable to estimate the number of ‘missed’ acute cases from backwards-projection of chronic cases. However, due to the substantial lag time (at least 10-30 years) between exposure and chronic symptoms, this was not possible with the data available from SINAN. However, should this topic be revisited in the following decades, a back-projection approach may be useful for retrospectively estimating the underreporting rate.

Our model uses Knorr-Held Type I spatio-temporal interactions *δ*, which are the least sophisticated of the structures outlined by Knorr-Held (2000). This strategy essentially estimates uncorrelated space-time fixed effects, which we found to be ideal for precise internal estimation; however, since these fixed effects lack a temporally autoregressive definition, it is not possible to use these fixed effects to project the model to predict future incidence without introducing meaningless statistical noise. In our prediction we simply dropped the interaction term from the model; in effect, only the un-interacted spatial and simulated random walk temporal terms were used for future projection. We believe that using the basic Type I interactions allowed for better estimation of the independent spatial and temporal components as we found that the Type IV term is only weakly identified in our model; so this decision was not without benefit. Choice of a spatio-temporal interaction that includes a temporally autoregressive term, such as Type II or Type IV (see supplementary material for more elaboration), would allow for projection in the form of a random walk through interaction space-time. For example, should a Type IV prior be used that assumes total spatial and temporal dependence in the interaction, future space-time interaction terms can be simulated as a random draw from a Multivariate Normal distribution with a variance-covariance matrix derived from the previously-estimated interaction terms, similar to the simulation procedure for spatially autocorrelated data (Banerjee, Carlin, and Gelfand 2015). Special care will likely be needed to assure propriety of the Type IV distribution, which is outside of the scope of the present study.

Our population projection would benefit from a more precise estimation methodology than the crude exponential growth model used here. Future analysis should consider municipality, year, and age-specific rates of fertility, mortality, and migration—especially internal migration—to inform population projections. We were unable to obtain these quantities at the level of spatial and temporal granularity required. We believe that in the short term—namely, the single decade between 2020 and 2030—this crude methodology allows for understanding how heterogeneity in growth rates may relate to future incidence of Acute Chagas Disease. Nonetheless, it is not suitable for long-term projections. Model-based estimates that utilize the readily-available state-level rates to determine small-area estimates, possibly similar to the Lee-Carter (1992) procedure or the one used by Alexander, Zagheni, and Barbieri 2017, may allow for more precise estimation of future municipality-level population. As well, an extension of the model to predict age-specific rates of Chagas disese may aide researchers in planning for interventions.

Finally, we make the critical assumption in both models that population and climate change are the sole drivers of future incidence. Other factors, such as housing construction materials, poverty, habitat destruction, and residential or industrial development encroaching on *Triatomine* habitats may increase affect rates of Chagas disease even in the absence of population or climate change. Further, the inherent assumption of linearity in our model assumes that as climate and population change, predicted incidence of Chagas disease will respond. This may not be the case: while climate may partially determine the geographic distribution of *Triatomines*, which may in turn affect incidence, it is likely that the relationship of climate with *Triatomine* populations is too complex to be captured by a linear model of the sort used here. Since there are many species of *Triatomines*, each with different habitats, behaviors, and virulence, the ultimate effect of climate on Chagas incidence is undoubtedly complex and nonlinear. Further, our climate model has a slightly higher error than the main model despite the inclusion of additional covariates, indicating that the model may be suffering from overfitting. As well, many exogenous factors could affect the distribution of *Triatomines* under future climate conditions, including interventional strategies to limit *Triatomine* habitats like the residential insecticidal campaigns of the 1980s, development of urbanization and infrastructure, and climate adaptations, environmental destruction and conservation practices that may affect *Triatomine habitats*. Should future climate conditions create new habitats for *Triatomines*, it is not clear at present if the insects are mobile enough to find these habitats, or if accidental importation by humans—such as improper handling of lumber—may catalyze a shift in *Triatomine* distribution.

### Replication Code

Replication code is publicly available at https://github.com/eroubenoff/chagas_modeling.

## Supplementary Material

### S1 Adjacency Matrices for Geostatistical Models

For the analysis of Chagas disease, we focus on methods for areal or polygon data, which refer to a region of space which contains a subset of the observations of interest. Polygonal data is a common format for administratively-collected spatial data, often representing a governmentally-defined area—such as a state or province equivalent, city or municipality, or even more specific form such as census tract or block. Areal data exist in contrast to point-referenced data, which instead link each observation with longitude and latitude coordinates. Whereas areal data can be generated from point data using a simple point-in-polygon operation, the reverse process is not possible as the specific coordinates are lost when points are tallied within polygons. Critical in any spatial statistcal work is the concept of the neighborhood matrix: a mathematical representation of geographic adjacency. For example, this 3x3 grid could be representing by binary neighborhood matrix W:

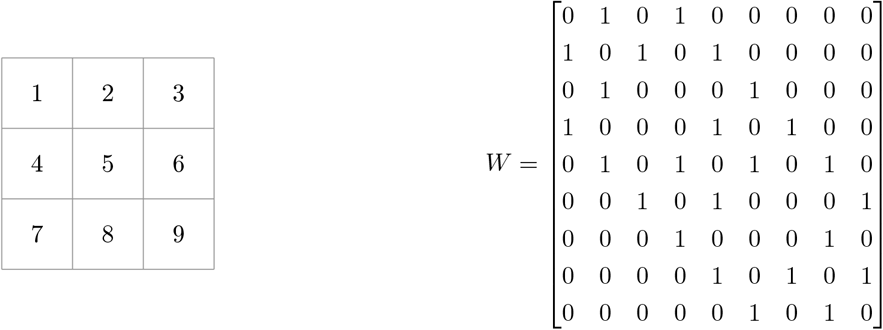

A symmetric binary matrix like this is most common for representing adjaceny, but can easily be extended to include reciprocal distance weights, higher-order neighbors, or measures of connectivity that are not strict adjacency (for example, consider transit networks or other transportational features that mean the travel time between two locations is not linear with distance). While estimates will change between different matrices W, the following distributional properties remain the same.

In the general case of eq. 1 with a non-binary neighborhood matrix ***W***, the equivalent, generalized model can be parameterized:

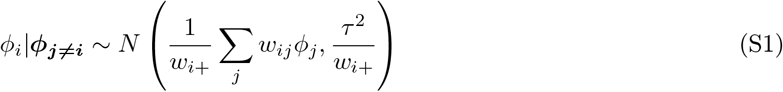

where *w*_*i*+_ =Σ_*j*_ *w*_*ij*_, or the sum of matrix ***W*** row *i*.

### S2 Commentary on the BYM-type model

In their 1991 paper, BYM use the IAR distribution (eq. 1) to fit a log-linear Poisson GLM of the form:

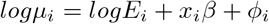

where *μ*_*i*_ is the rate of disease occurrence in unit *i, E* is expected count unit *i, x* and *β* are explanatory variables and regression coefficients, and error term *ϕ*_*i*_ has the prior distribution of eq. 1. This model, which utilizes internal standardization, was called by Banerjee, Carlin, and Gelfand “cheating (or at least ‘Empirical Bayes’)” since the *E*_*i*_ are not fixed but rather itself part of the data, posing a ‘null hypothesis’ about if there were to be absent a spatial pattern (BCG, p.151)^6^. As well, BCG and Leroux et al (2000) note that this model may have poor performance as using the CAR prior alone as an error term may over-smooth aspatial variation, which may be mechanistically important to the model.

The IAR distribution is conditionally specified for each geographic unit and is improper, meaning that it does not integrate to produce a valid probability distribution, instead only able to show the proportional density between spatial units. This is problematic for stochastic generation and maximum likelihood estimation, but is valid for Bayesian inference as posterior density need only be proportional to the prior density (Besag, York, and Mollie 1991). However, it is possible and mathematically convenient to consider equation 1 in its joint, albeit improper, form. Besag (1974) showed that fully conditional distributions of this type can utilize Brook’s Lemma (1964) to recover the full conditional form, as a multivariate normal distribution with mean 0 and variance-covariance matrix related to the adjacency matrix. Banerjee, Carlin, and Gelfand demonstrate this concisely, determining the joint distribution of *ϕ* from a fully specified set of conditionals:

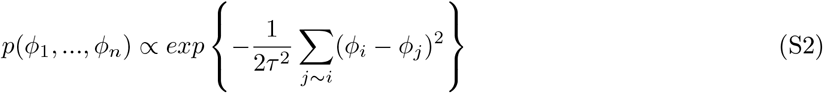

which is also known as the pairwise-differences formula (Banerjee, Carlin, and Gelfand 2015, p.81, eqn. 4.16). Equation S2 can be utilized to provide convenient estimation in Bayesian MCMC, using software such as Stan as pairs of adjacent units can be efficiently stored in program memory, and the proportional density can be quickly computed without the need for matrix inversion. Since Stan estimates the proportional log-density of *ϕ* up to a constant, Morris et al. 2019 demonstrate that equation S2 can be quickly evaluated as:

~~~
     phi ∼ sigma * -0.5 * dot_self(phi[node1] - phi[node2]);
     sum(phi) ∼ normal(0, 0.001 * N);
~~~

where `sigma` is the precision rather than the variance, where `node1` and `node2` are vectors of adjacent pairs, `dot_self` takes the dot product of the vector with itself, and the second line indicates that `phi` is subject to a soft sum-to-0 constraint. Due to this computational efficiency and problematic assumptions needed to make this distribution properly integrate, BCG recommend that the IAR model be used only in the case of a Bayesian prior, and may be frequently the optimal choice for geostatistical inference (BCG, ch. 4 and BCG, ch. 6, p.155)

The expression in S2 is an improper probability distribution since the joint probability density is only proportional to the derived expression. This is because the variance-covariance matrix implied is singular, meaning the inverse does not have a unique solution and as a result the distribution does not necessarily sum to one, as required for valid probability distributions. For a non-mathematical explaination, consider that each observation is entirely dependent on its neighbors, which allows us to estimate the total distribution only on relative terms without a ‘ground truth’ or some external source centering the distribution. To demonstrate this impropriety, BCG (p.81) derive equation 1 by beginning with adjacency matrix ***W***, which has *w*_*ij*_ = 1 if *i* and *j* are neighbors and 0 otherwise; matrix ***B*** where *b*_*ij*_ = *w*_*ij*_*/w*_*i*+_, or a row-standardized version of matrix ***W*** ; and ***D***, a diagonal matrix where *d*_*ii*_ is equal to the number of neighbors of *i* and 0 otherwise. Then, equation 1 can be written in the conditional form as 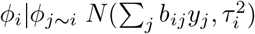 since ***B*** is the row-standardized version of ***W*** . This would imply that ***ϕ****∼MVN*(**0**, [*τ*(***D****−****W***)]^*−*1^). Temporarily disregarding *τ*, calculating the covariance matrix Σ^*−*1^ of this distribution involves the calculation (***D*** *−* ***W***)**1** = **0**, which is singular; effectively, too many variables without a constraint to preserve propriety (BCG, p.81). It is possible to make this distribution proper with an additional parameter, often denoted *ρ* (*α* in Morris et al. 2019), by defining the inverse covariance matrix Σ^*−*1^ = ***D*** *−ρ****W***, so long as *ρ* is chosen to find a singular solution. BCG list the bounds under which *ρ* will provide a non-singular solution, which is related to the eigenvalues of matrices ***D*** and ***W***. Then, the full conditional distribution becomes:

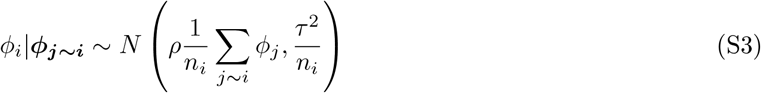

BCG write that *ρ* is sometimes reported as being the degree of spatial autocorrelation, but it is clear from equation S3 that the resultant expression rather expresses some *proportion* of the spatial gaussian process (p.82). As well, *ρ* does not map clearly onto any other measures of spatial autocorrelation, like Moran’s I or Geary’s C, and thus its interpretation outside of the model is limited.

Further, the authors remark that the proper CAR model may be attractive in cases where the spatial pattern is weak, and the improper CAR model may over-smooth heterogeneity. However, in simulation, the proper CAR model has been shown to nearly always converge on values of *ρ* close to 1, as when *ρ* is less than 1 there presents an identification challenge between the spatial random effects and the non-spatial random effect. BCG remark (p.155) that it appears that the data always *want ρ* to be close to 1. In conclusion, BCG recommend that the improper IAR model be only used as a Bayesian prior, or in the frequentist case, use of a SAR or other proper probability distribution.

As noted above, two issues present with using the IAR model alone as a prior for a spatially autocorrelated error term. First, the IAR model is known to show poor performance when spatial autocorrelation is not very strong, otherwise it will oversmooth random variation in the data. This issue is rectified with a proper CAR model, but as noted above, BCG do not recommend usage of the proper CAR prior. Second, the IAR variance parameter *τ* has an ambiguous function, and sources differ as to its interpretation. While Leroux (2000) states that this parameter represents both autocorrelation and over-dispersion simultaneously, but Banerjee, Carlin, and Gelfand write that this parameter should not be taken as representing spatial autocorrelation in any mechanistic way.

Ideally, we would have included the BYM2-type spatial convolution term in both the Bernoulli and Poisson processes, however in model development we were unable to reach convergence with the model specified as such: the convolved spatial error for the Poisson parameter *λ* failed to be identified. Recall that this term is specified in both the Bernoulli and Poisson parameters as 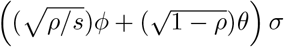, where *ϕ* is the IAR model, *θ* is *N* (0, 1), *ρ* represents the proportion of variance having a spatial pattern rather than unstructured error, and *σ* is the overall variance. The convolved spatial process in the Bernoulli part showed excellent convergence and mixing with posterior estimates of *ρ*_*π*_ *≈* 0.75. As a result, both the spatially clustering term *ϕ* and the spatial heterogeneity term *θ* contributed to posterior estimates of πand the likelihood of the model. However, in the Poisson part, median posterior estimates of *ρ*_*λ*_ *≈* 0 such that the convolved error term *≈ θσ*. As a result, the sampler could not identify values for *ϕ*_*λ*_, functionally searching the entire parameter space for *ϕ*_*λ*_ without any effect on the likelihood of the model. Ultimately, this caused slow evaluation and divergences, but did not affect the resultant values of other parameters.

### S3 Zero-Inflated Models and their efficient estimation in Stan

To assess the appropriateness of the ZIP distribution for our data, we conducted a naive maximum likelihood estimate of the Poisson parameter *λ* and the Zero-Inflated Poisson parameters π, *λ* without any adjustment for spatial structure, annual deviations, or covariates. Likelihood functions for both the Poisson-only specification and ZIP specification were optimized over the municipality-year counts of Chagas incidence using R’s multivariate `optim` routine. The maximum likelihood estimate of Poisson-only parameter *λ* was 0.1053 with log-likelihood *ℓℓ* = 57773.01, and the estimate of ZIP parameters were *π* = 0.9849 and *λ* = 6.9885 with *ℓℓ* = 19330.45. To test the relative fit of both models, we conduct Wilk’s test for likelihood ratios, which assumes that the ratio of two likelihoods is asymptotically distributed as *χ* ^2^(*df* = *df*_*H*1_ *− df*_*H*0_). Taking the Poisson-only specification as the null hypothesis and the ZIP specification as the alternative, the probability of observing these data generated by the Poisson-only specification instead of the ZIP specification is *p ≈* 0. Hence, we can comfortably reject the Poisson-specification in favor of the ZIP specification.

Using ZIP models may provide an additional computational advantage over a regular Poisson specification. We found that a model of the type laid out in the previous sections, which models the count of Chagas disease as a Poisson-distributed GLM with terms for fixed effects for spatial and aspatially-clustered errors, showed slow evaluation and poor estimation. While the sheer dimensionality of the model—approximately estimating 6 parameters for 5000 municipalities across 19 years—was undoubtedly responsible for part of the problem, we hypothesized that the complicated posterior geometry caused by the overdispersion of 0s in the dataset was partially to blame. To test this hypothesis, we ran two test cases each with a single UF over the first two years of the study period. We chose Pará (PA), which has the highest number of Chagas cases at 5259 over the 19 year study period in 143 municipalities, and Roraima (RR), with the second lowest number of cases at 10 cases in 15 municipalities^7^. In Stan, the test models were run for 500 warmup iterations and 500 sampling iterations. For PA, the model completed evaluation in 2508 seconds for an average parameter effective sample size (ESS) of 2234.738 (SD = 1413). However, despite RR having one tenth the number of municipalities of PA, the model for RR took more than twice as long to evaluate—5994.4 seconds— for an average parameter ESS of 1416.437 (SD = 451.5). Both models showed convergence 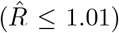for more than 99% of parameter estimates. When this test was replicated using the ZIP model with an autoregressive component in the Bernoulli part only, the PA model evaluated in 46.2 seconds with an average parameter effective sample size of 470.96, and the RR model evaluated in 57 seconds with an average ESS of 25.39.

Naive implementation of Spatio-Temporal statistical models involves many pairwise comparisons, which can be prohibitively computationally expensive for MCMC estimation. For example, our spatio-temporal adjacency structure may contain not only neighboring observations between all 5000 municipalities in Brazil, but the temporally-correlated neighbors as well. Assuming that an average municipality has 4 spatial neigh-bors and 4 temporal neighbors, this will result in inverting a neighborhood matrix of over 1.6 billion elements which is not reasonably evaluated with standard computing resources, let alone for thousands of MCMC iterations. We have taken many steps towards quick and efficient estimation in lieu of this challenge, which is a major contribution of this research in addition to the primary substantive estimation of Acute Chagas Disease incidence.

When sampling a ZIP GLM in Bayesian software such as Stan, we will have to write a custom log-probability mass function (LPMF, in Stan terms) to cover zero-inflation. First, we assume that Bernoulli parameter π is estimated on a logit scale and Poisson parameter *λ* is estimated on a log scale, as is common for GLMs. Second, we must evaluate the probability density on the log scale. The following is an unoptimized ZIP estimation, adapted from Stan’s documentation for Zero-Inflated Poisson models^8^:

~~~
for (t in 1:T){
    for (n in 1:N) {
        if (y[t,n] == 0) {
         target += log_sum_exp(bernoulli_logit_lpmf(1 | pi[t,n]),
                               bernoulli_logit_lpmf(0 | pi[t,n])
                               + poisson_log_lpmf(y[t,n] | pi[t,n]));
        } else {
          += bernoulli_logit_lpmf(0 | pi[t,n])
                      + poisson_log_lpmf(y[t,n] | pi[t,b]);
        }
    }
}
~~~

where `target` is the log-probability of the model and `log_sum_exp(a,b)` = log(exp(a) + (exp(b)). Clearly, this is a highly inefficient way to evaluate the model since the log-probability statement is conditioned on the data `y` which is known and constant through the course of the simulation. In computational efficiency terms, each evaluation of the likelihood will complete in *O*(*T · N*) time, meaning that the time to evaluate the log-probability statement is proportional to the number of municipalities times the number of years. For our application to Chagas Disease in Brazil, which contains observations of approximately 5000 municipalities over 19 years, this becomes extraordinarily slow, evaluating 1000 warmup and sampling iterations on the scale of 12-24 hours.

To optimize this Stan modeling statement, the Stan documentation recommends partitioning the data into zero and non-zero elements and evaluating them separately, but does not elaborate on how to do so in a GLM framework, which we have developed for our application. Doing so will allow for separate, efficient vectorized evaluation of the Bernoulli and Poisson GLM statements. Indeed, as explained elsewhere, vectorization is one of the primary benefits of using Stan over other Bayesian MCMC software suites, since vectorized probability statements evaluate much faster and with less overhead than doubly-looped functions. First, consider a matrix of counts *Y* with dimensions *T* (number of years) and *N* (number of municipalities). From *Y*, we will derive two matrices `zero_idx` and `nonzero_idx` with the same dimensions, containing the indices of zero and nonzero observations, and supported by vectors `zero_max` and `nonzero_max` with dimension *T*, where each element contains the annual number of zero and nonzero entries. In this way, for matrix row *t ∈ T* columns [1 : `zero_max[t]`] contains the index of municipalities with zero entries, and [`zero_max[t]` + 1 : *N*] are uninitialized. Beginning with matrix of counts *Y* :

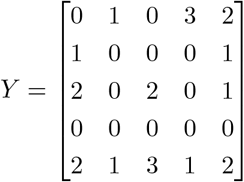

that yields derived zero and non-zero matrices:

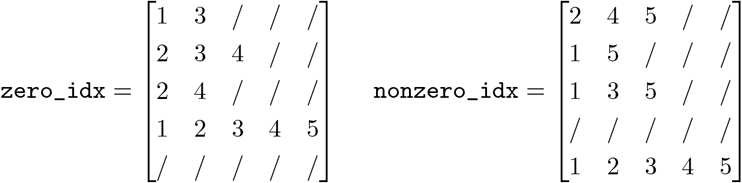

with max vectors:

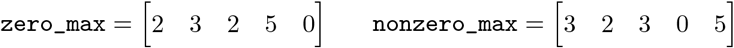

In Stan, zero-counts in year *t* can be be easily indexed from *Y* as `Y[t, zero_idx[t, 1:zero_max[t]]]`and nonzeros as `Y[t, nonzero_idx[t, 1:nonzero_max[t]]]`. Essentially, these sparse matrices are an efficient way to store ragged arrays, which are not supported natively in a C-based language like Stan. The R code for generating these matrices and vectors from matrix *Y* in:

~~~
n_T = nrow(Y)
N = ncol(Y)
zero_max = array(rep(0,n_T))
nonzero_max = array(rep(0,n_T))
zero_idx = matrix(0, nrow = n_T, ncol = N)
nonzero_idx = matrix(0, nrow = n_T, ncol = N)
for (t in 1:n_T) {
  for (n in 1:N){
   if (Y[t,n] == 0) {
     zero_max[t] = zero_max[t] + 1
     zero_idx[t, zero_max[t]] = n
   }
   else {
     nonzero_max[t] = nonzero_max[t] + 1
     nonzero_idx[t, nonzero_max[t]] = n
   }
  }
}
~~~

Then, we can turn to writing a log probability mass function describing equations 7 and 8 that takes advantage of this vectorization. First, recall that π is on the logit scale and *λ* is on the log scale, and we wish to evaluate the probability on the log scale. Assuming that π and *λ* have been transformed using their corresponding inverse-link functions, this yields likelihood:

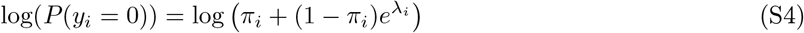

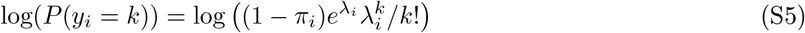

Equation S4 is problematic in that it does not simplify to built-in Stan probability statements, but can be written in a way that is efficiently vectorized^9^. Luckily, equation S5 simply evaluates to two independent expressions of Bernoulli probability and Poisson probability on the log scale, respectively. This means they can be evaluated in Stan as:

~~~
vector[N] pi_inv_logit;
vector[N] lambda_exp;
for (t in 1:T) {
    pi_inv_logit = inv_logit(pi[t]);
    lambda_exp = exp(lambda[t]);
    // Zeros
    target += sum(log(
           pi_inv_logit[zero_idx[t, 1:zero_max[t]]] +
        (1-pi_inv_logit[zero_idx[t, 1:zero_max[t]]]) .*
        exp(-lambda_exp[zero_idx[t, 1:zero_max[t]]])
      ));
     // Nonzeros
     target += bernoulli_lpmf(
                rep_array(0, nonzero_max[t]) |
                pi_inv_logit[nonzero_idx[t,1:nonzero_max[t]]]
          ) + poisson_lpmf(
                y[t, nonzero_idx[t, 1:nonzero_max[t]]] |
                lambda_exp[nonzero_idx[t, 1:nonzero_max[t]]]
          );
}
~~~

In a test case, this vectorized model evaluated 100 warmup iterations and 100 sampling iterations in 1099 seconds, more than 10 times faster than the non-vectorized example.

### S4 Knorr-Held Spatio-Temporal Models

The other priors outlined in Knorr-Held (2000) are, respectively:

- Type I interaction, where all interaction terms are *a priori* independent:

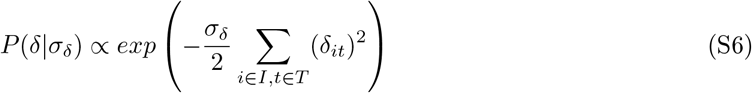

which is suitable if the space-time interaction does not have any structure.
- Type II interaction, where each spatial unit follows a 1st order random walk independent of its neighbors:

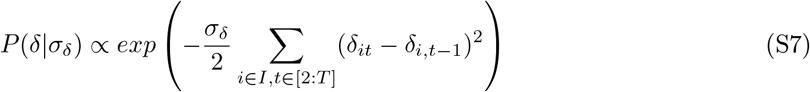

which is suitable if temporal trends differ between spatial units and the temporal trends do not have any structure in space.
- Type III interaction, where interaction effects follow and intrinsic autoregression such as the type laid out in equation 1, but are indepdent at each time:

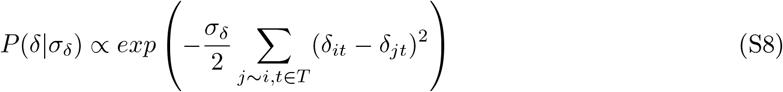

which is suitable if the spatial trends differ between time points, but the temporal trends do not have any structure in space.
- Type IV interaction, perhaps the most methodologically and conceptually interesting, where effects are totally dependent over space and time:

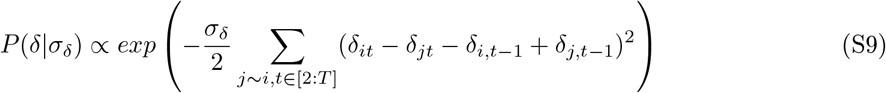

Which defines a space-time Markov random field and is suitable if temporal trends are different from location to location but are more likely to be similar in adjacent locations. This prior can be written in Stan as:

~~~
real knorr_held_type4_lpdf(vector delta_t, vector delta_tm1, int N, int[] node1, int[] node2) {
    return -0.5 * dot_self(delta_t[node1] - delta_t[node2] - delta_tm1[node1] + delta_tm1[node2]) +
             normal_lpdf(sum(delta_t) | 0, 0.001*N) ;
}
~~~

where `delta_t` is the value of *δ* at time *t*, `delta_tm1` is the value of *δ* at time *t −* 1, `node1` and `node2` indicate adjacent pairs of nodes, and the `normal_lpdf` statement indicates a soft sum-to-0 constraints for *δ*_*t*_, as done for the ICAR prior above.

At face value, interaction Type IV would be the most useful for our purposes, however in model development we found that this model both was under-identified and over-smoothed random variation in the data. Instead, we opt for Type I priors, which are both simpler to estimate and more easily identified. Theoretically, type IV interactions are comparing not only the first degree neighbors—each observation with its spatial neighbors and previous observation—but also the 2nd order neighbors—the spatial neighbors of temporal neighbors, or equivalently, the temporal neighbors of spatial neighbors (Knorr-Held 2000). Essentially, this prior is an extension of the pairwise-differences CAR prior (eq S2) to the temporal dimension. Where the pairwise CAR prior focuses on the differences between adjacent units, the Knorr-Held Type IV prior includes the differences between adjacent units in the current time period and the prior time period. Knorr-Held remark that such a model may be useful for modeling the spatio-temporal spread of both infectious diseases and non-infectious diseases where the underlying risk has a spatio-temporal pattern, as is appropriate for our application to Chagas disease. For the first time point *t* = 1, the ‘previous’ time period *t* = 0 is unavailable, so for this case only the ‘previous’ time period is instead taken to be a 0 vector, at which point the model simplifies to the ICAR prior. If the full model is specified as in equation 5, it is likely that identifiability will be poor without highly informative priors, as was the case for the BYM model above. For the present application to Chagas disease, it may be possible to disregard the time trends *α* and *γ*, since most locations begin and remain absent Chagas disease throughout the duration of study.

### S5 Stan model code, edited slightly for clarity

~~~
functions {
  real icar_normal_lpdf(vector phi, int N, array[] int node1, array[] int node2) {
    // Soft sum-to-zero constraint
    return -0.5 * dot_self(phi[node1] - phi[node2]) + normal_lpdf(sum(phi) | 0, 0.001*N);
  }
}
data {
    // Number of municipalities
    int<lower=0> N;
    // Number of years
    int<lower=0> T;
    // Number of adjacent edges
    int<lower=0> N_edges;
    // node1[i] adjacent to node2[i]
    array[N_edges] int<lower=1, upper=N> node1;
    // and node1[i] < node2[i]
    array[N_edges] int<lower=1, upper=N> node2;
    // count outcomes
    array[T,N] int y;
    // Population exposure
    array[T,N] int E;
    // Scaling factor-- scales variance of spatial effects
    real<lower=0> scaling_factor;
    // indices of zero counts
    array[T,N] int zero_idx;
    // Max number of zero counts
    array[T] int zero_max;
    // indices of nonzero counts
    array[T,N] int nonzero_idx;
    // max number of nonzero counts
    array[T] int nonzero_max;
}
transformed data {
    // Logged population
    array[T] vector[N] log_E;
    for (t in 1:T) {
      log_E[t] = to_vector(log(E[t,1:N]));
    }
}
parameters {
    // Bernoulli part: Knorr-Held model
    // Intercept real mu_pi; real mu_lambda;
    // Structured temporal trend
    vector[T] alpha_pi;
    vector[T] alpha_lambda;
    real<lower=1e-10, upper=10> sigma_alpha_pi;
    real<lower=1e-10, upper=10> sigma_alpha_lambda;
    // Structured spatial pattern vector[N] phi_pi;
    // vector[N] phi_lambda;
    // Unstructured spatial pattern
    vector[N] theta_pi;
    vector[N] theta_lambda;
    // Proportion of spatial/aspatial error
    real<lower=0, upper=1> rho_pi;
    // real<lower=0, upper=1> rho_lambda;
    real<lower=1e-10, upper=10> sigma_convolved_pi;
    real<lower=1e-10, upper=10> sigma_convolved_lambda;
   // Knorr-Held Type I spatio-temporal interaction
    array[T] vector[N] delta_pi;
    array[T] vector[N] delta_lambda;
    real<lower=1e-10, upper=10> sigma_delta_lambda;
    real<lower=1e-10, upper=10> sigma_delta_pi;
}
transformed parameters{
    array[T] vector[N] pi; // Bernoulli GLM term
    array[T] vector[N] lambda; // Poisson GLM term
   for (t in 1:T) {
      pi[t] = inv_logit(mu_pi +
              alpha_pi[t] +
              sigma_convolved_pi * (
                sqrt(rho_pi/scaling_factor) * phi_pi + sqrt(1-rho_pi)*theta_pi
              ) +
              sigma_delta_pi * delta_pi[t]);
      lambda[t] = exp(log_E[t] + mu_lambda +
                alpha_lambda[t] +
                sigma_convolved_lambda * (
                  // sqrt(rho_lambda/scaling_factor) *
                  // phi_lambda theta_lambda
                  // sqrt(1-rho_lambda)*theta_lambda
                ) +
                sigma_delta_lambda * delta_lambda[t]);
    }
}
model {
    // Intercepts
    mu_pi ∼ normal(-10, 10);
    mu_lambda ∼ normal(-5, 10);
    // Structured temporal trend
    alpha_pi[1] ∼ normal(0, sigma_alpha_pi);
    alpha_pi[2:T] ∼ normal(alpha_pi[1:(T-1)], sigma_alpha_pi);
    sigma_alpha_pi ∼ gamma(2, 1);
    alpha_lambda[1] ∼ normal(0, sigma_alpha_lambda);
    alpha_lambda[2:T] ∼ normal(alpha_lambda[1:(T-1)], sigma_alpha_lambda);
    sigma_alpha_lambda ∼ gamma(2, 1);
    // Structured spatial patten
    phi_pi ∼ icar_normal(N, node1, node2);
    // phi_lambda ∼ icar_normal(N, node1, node2);
    // Unstructured spatial error
    theta_pi ∼ std_normal();
    theta_lambda ∼ std_normal();
    // Prior on Rho
    rho_pi ∼ beta(.5, .5);
    // rho_lambda ∼ beta(.5, .5);
    // Convolved variance
    sigma_convolved_pi ∼ gamma(2,1);
    sigma_convolved_lambda ∼ gamma(2,1);
    for (t in 1:T){
      // Interaction
      delta_pi[t] ∼ std_normal();
      delta_lambda[t] ∼ std_normal();
    }
    sigma_delta_pi ∼ gamma(2, 1);
    sigma_delta_lambda ∼ gamma(2, 1);
    // Likelihood
    for (t in 1:T) {
      // Vectorized ZIP
      // Zeros
      if (zero_max[t] > 0) {
        target += log(
               pi[t, zero_idx[t, 1:zero_max[t]]] +
           (1 - pi[t, zero_idx[t, 1:zero_max[t]]]) .*
           exp(-lambda[t, zero_idx[t, 1:zero_max[t]]])
        );
      }
      // Nonzeros
      if (nonzero_max[t] > 0) {
       target += bernoulli_lpmf(
                  rep_array(0, nonzero_max[t]) |
                  pi[t, nonzero_idx[t,1:nonzero_max[t]]]
             );
      target += poisson_lpmf(
                  y[t, nonzero_idx[t, 1:nonzero_max[t]]] |
                  lambda[t, nonzero_idx[t, 1:nonzero_max[t]]]
             );
      }
    }
}
~~~

**Table S1:** MCMC convergence diagnostics for main smoothing model selected parameters.

| Parameter | $\hat{R}$ 5th Quantile | $\hat{R}$ 95th Quantile | ESS 5th Quantile | ESS 95th Quantile |
| --- | --- | --- | --- | --- |
| $\pi$ | 1.00 | 1.01 | 941.00 | 3,708.96 |
| $\lambda$ | 1.00 | 1.00 | 6,116.68 | 10,733.15 |
| $\phi_\pi$ | 1.00 | 1.00 | 993.54 | 3,845.31 |
| $\theta_\pi$ | 1.00 | 1.00 | 8,329.99 | 12,173.52 |
| $\theta_\lambda$ | 1.00 | 1.00 | 4,293.10 | 11,373.93 |
| $\alpha_\pi$ | 1.00 | 1.01 | 430.58 | 635.58 |
| $\alpha_\lambda$ | 1.00 | 1.01 | 364.32 | 537.66 |
| $\delta_\pi$ | 1.03 | 1.03 | 316.20 | 316.20 |
| $\delta_\lambda$ | 1.00 | 1.00 | 734.21 | 734.21 |

**Figure S1:**
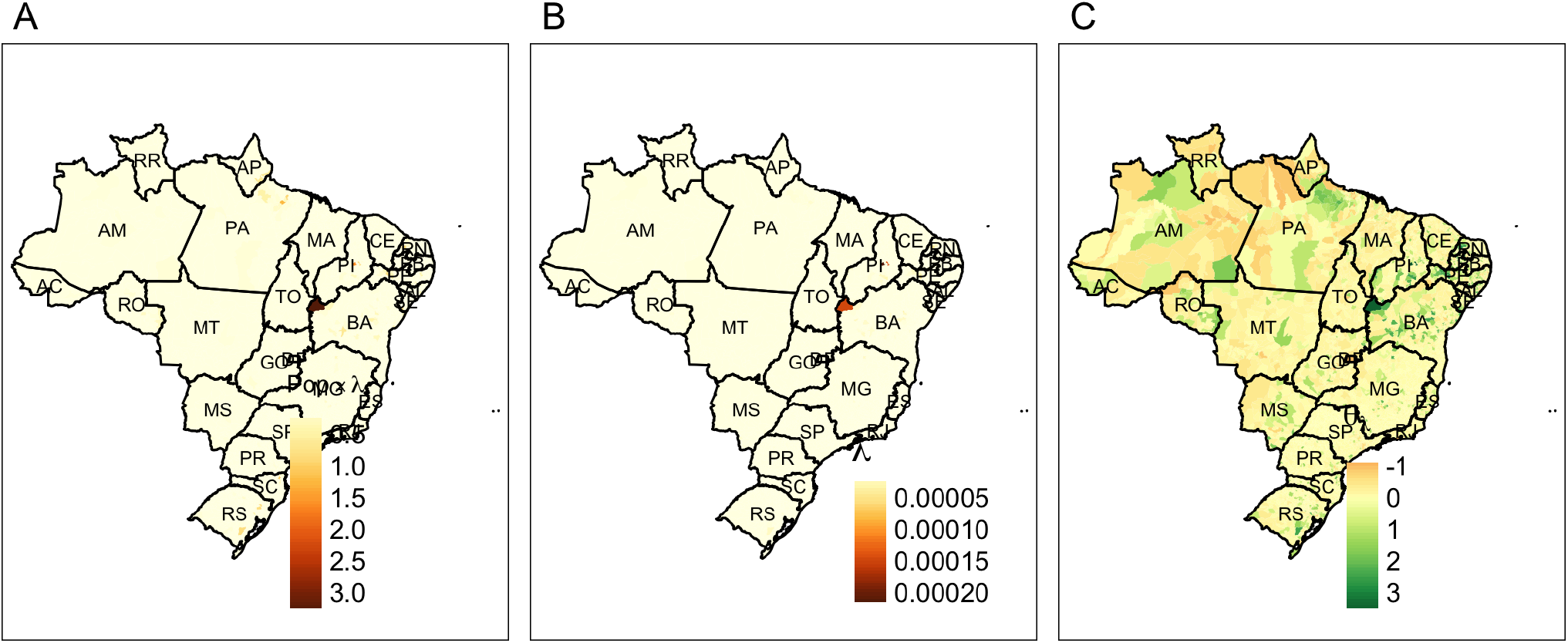
Spatial process in the Poisson term, without temporal effects. A: overall rate of Chagas, calculated as *Pop*_*i*_ *×λ*_*i*_, where *λ*_*i*_ = *exp*(*μ*_*λ*_ + *θ*_*i*_*\*σ*_*i*_); B: per-capita rate of Chagas *λ*, net of population; C: spatial heterogeneity term *θ*_*λ*_, with *N* (0, 1) prior.

**Figure S2:**
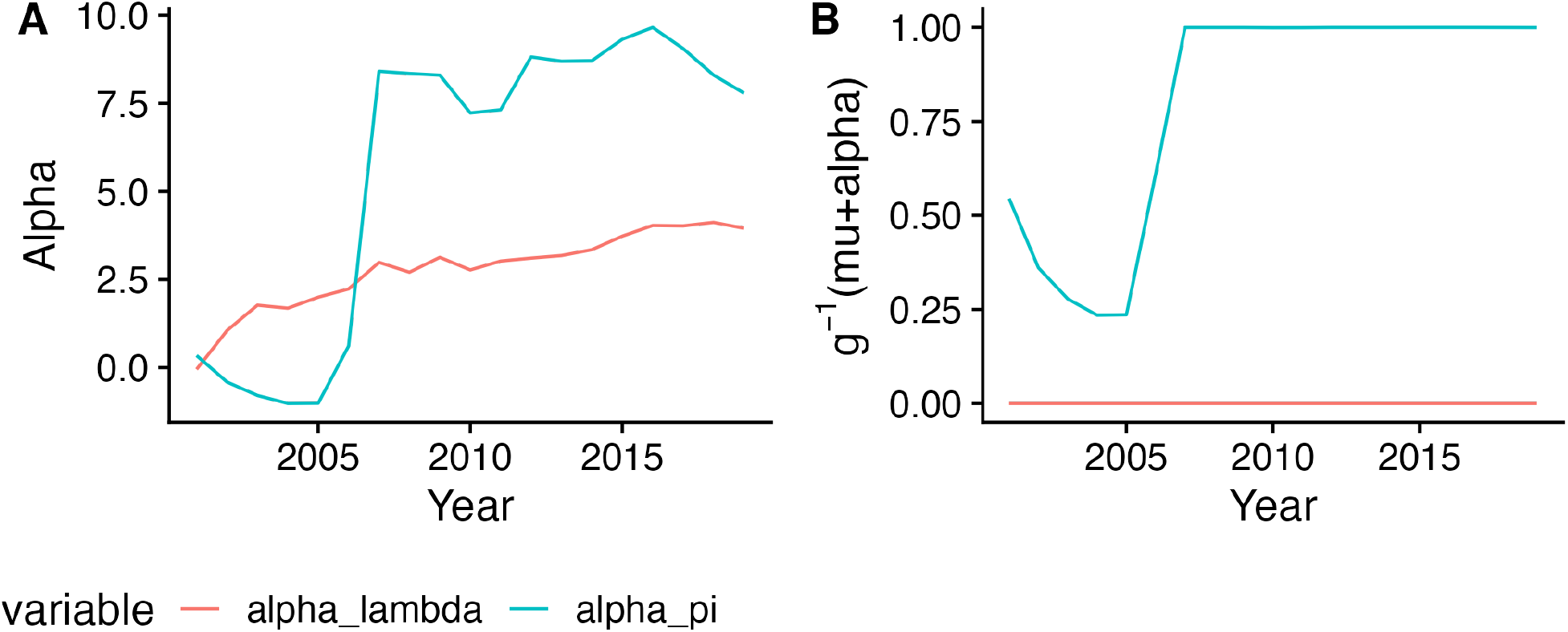
Global AR(1) time trend for Bernoulli and Poisson processes, on the (A) crude scale and (B) transformed scale, where the transformed scale is *logit*^*−*1^(*μ*_*λ*_ + *α*_*π*_) for the Bernoulli probability and *exp*(*μ*_*λ*_ + *α*_*λ*_) for the Poisson process. While the Poisson process always stays near 0, indicating that the rate of Chagas conditional on its presence in an area is stable over time, the global temporal trend of the Bernoulli parameter indicating probability of non-exposure drops initially, recovering to 100% by 2008. This implies that over the period of study, Chagas disease became much less global and more local in presentation.

#### S5.1 Main Model Additional Figures

#### S5.2 Climate Model Additional Figures

**Table S2:**
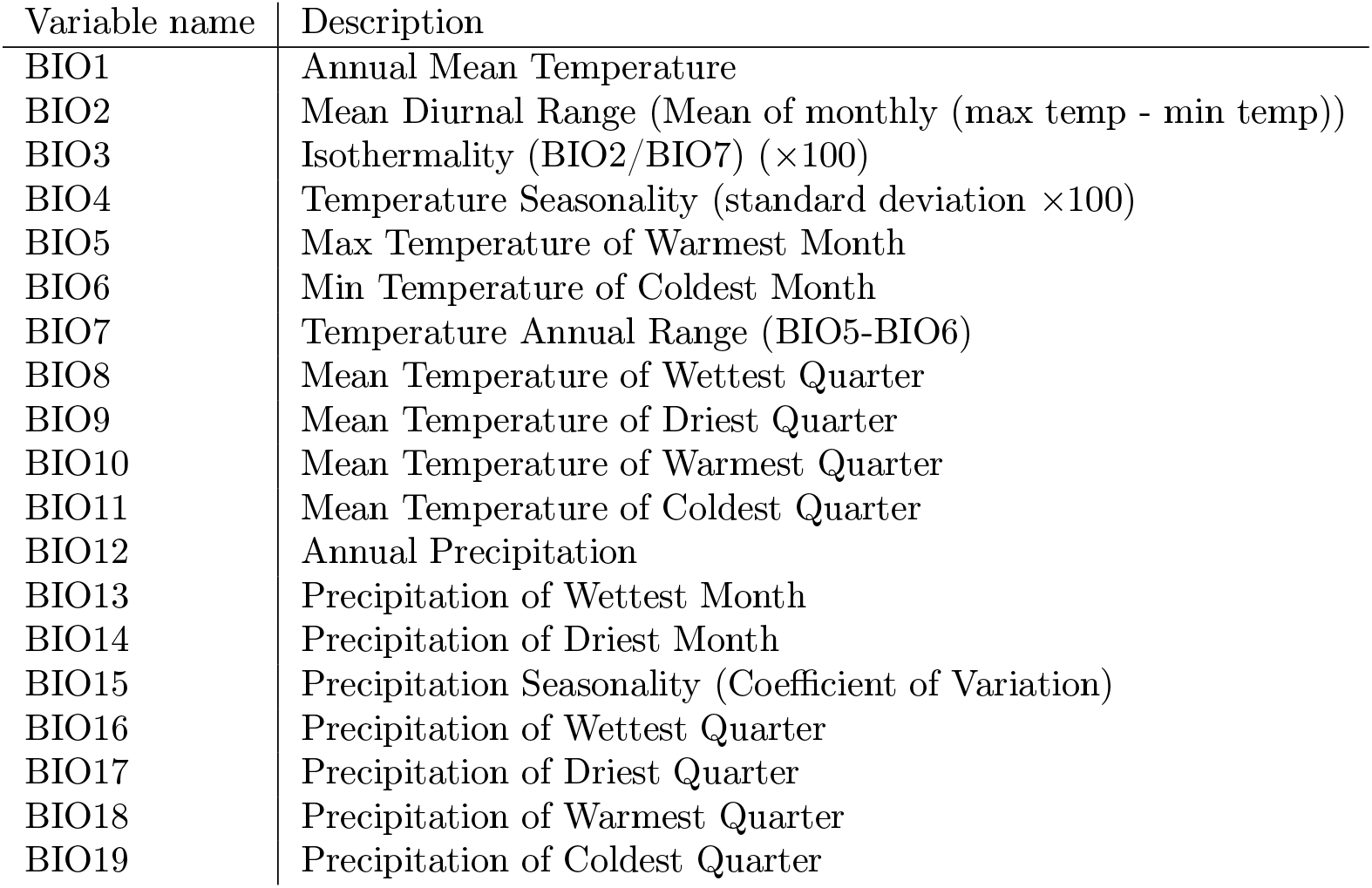
WorldClim suite of Bioclimatic variables downloaded from the Copernicus Climate Change Service’s Global Bioclimatic Indicators from 1950-2100 Derived from Climate Projections dataset.

**Table S3:**
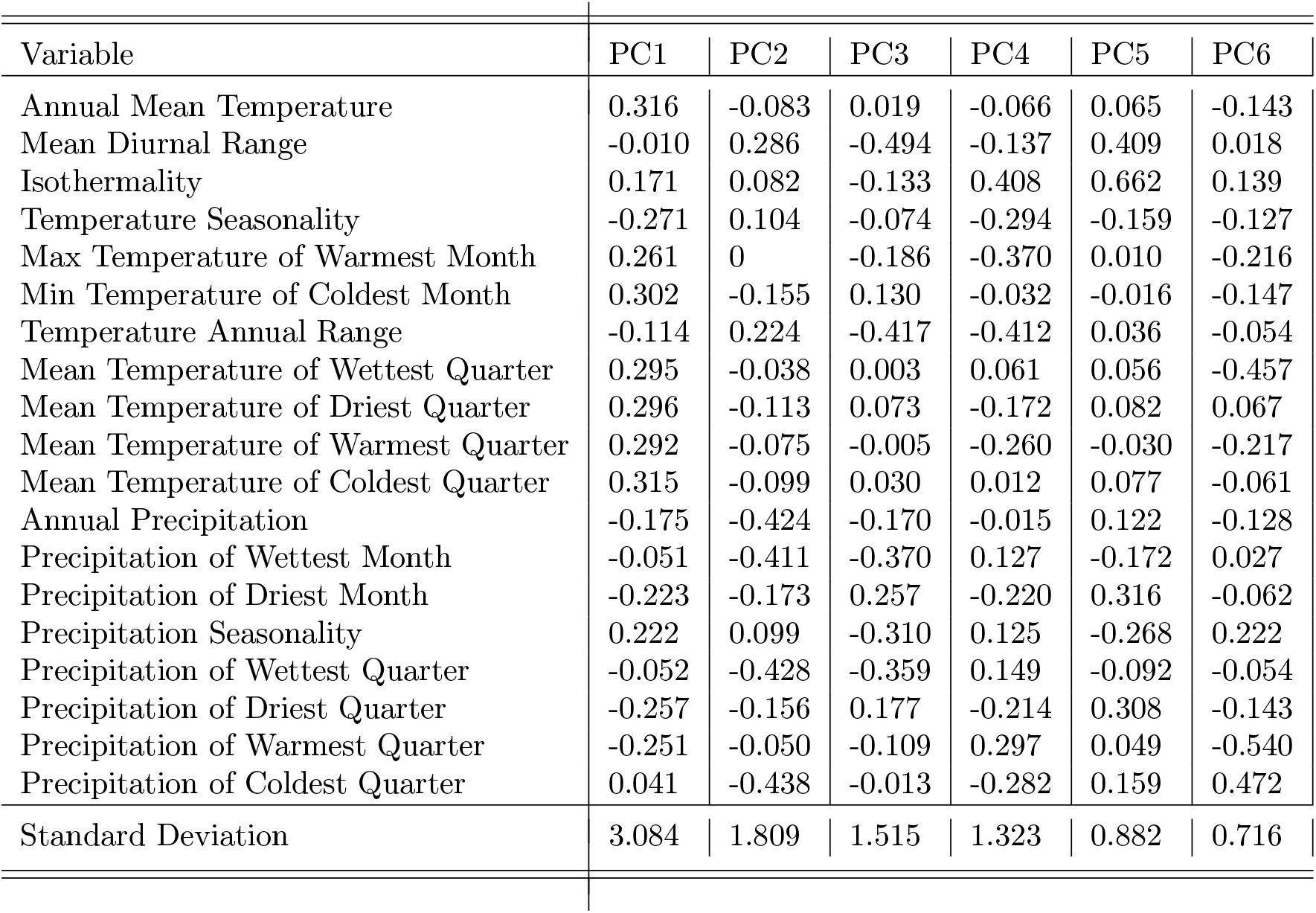
Principal Components 1-6 of the 19 WorldClim Bioclimatic Variables for median municpality-years in Brazil, 2000-2019.

**Figure S3:**
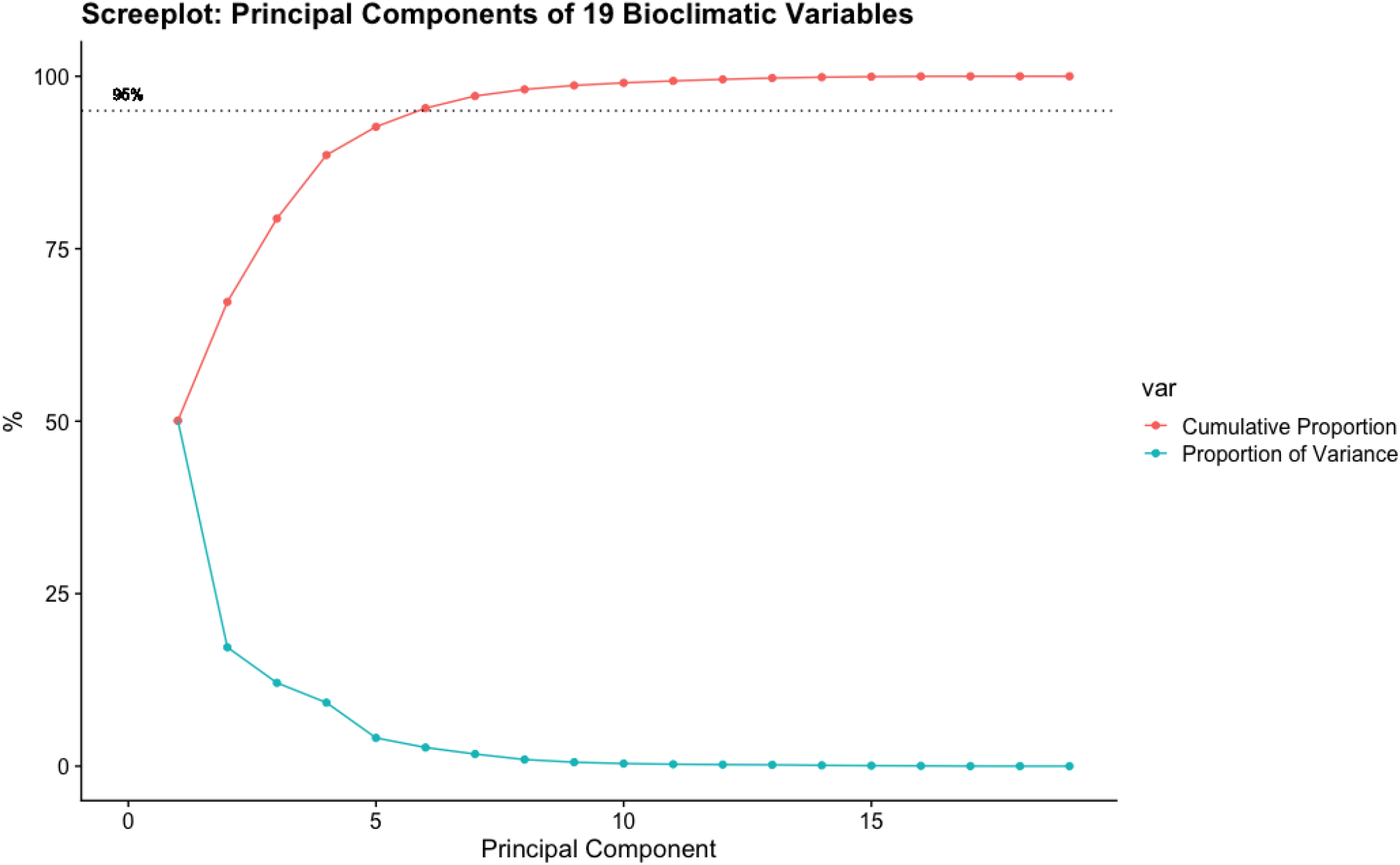
Screeplot of variance and cumulative variance explained by the first *n* principal components, with 95% of cumulative variance indicated by the dotted line.

**Figure S4:**
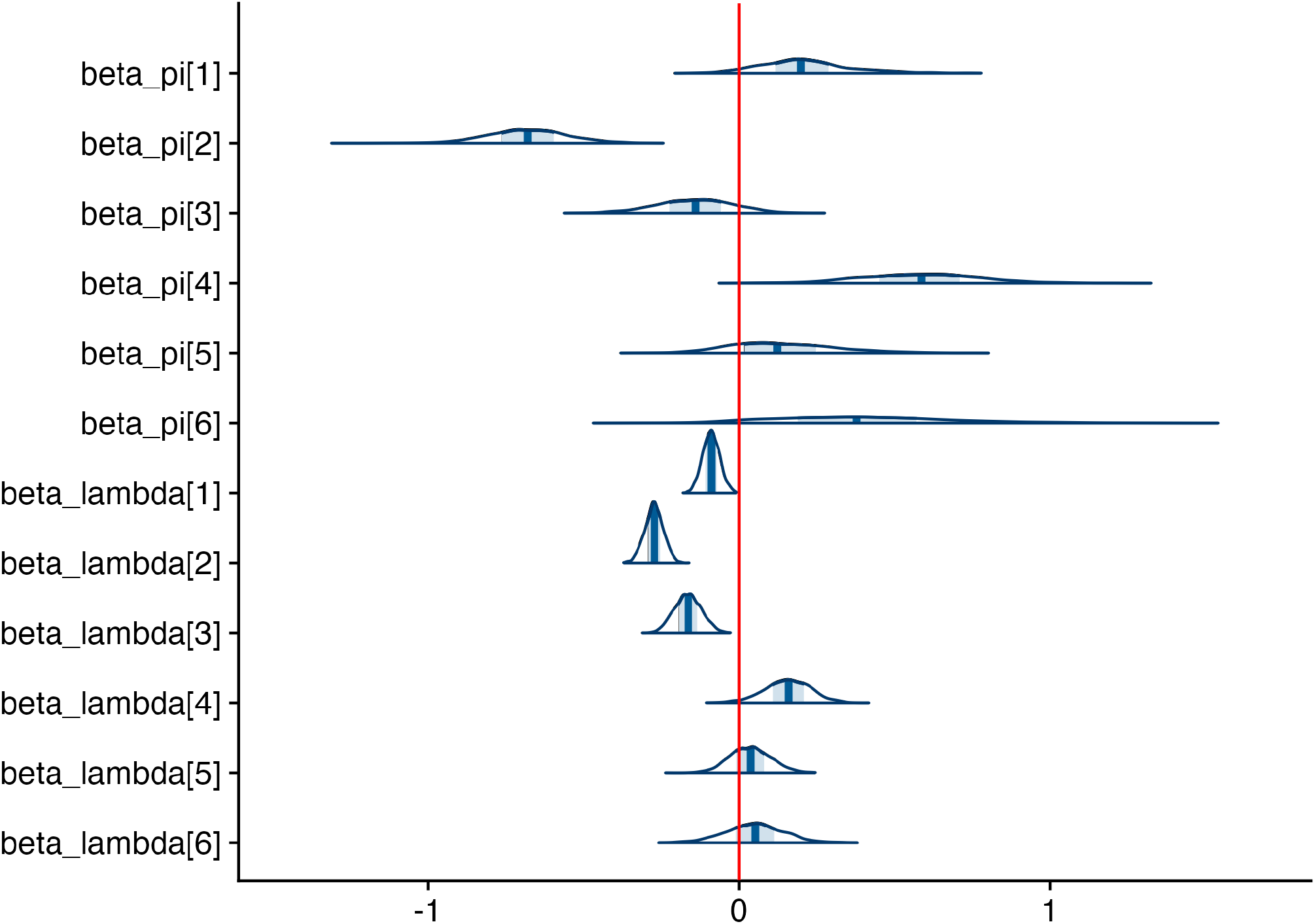
Plot of Climate Covariates, specified for both the Bernoulli process (π) and Poisson process (*λ*) as each municipality-year’s location in principal component space of the 19 WorldClim Bioclimatic Variables.

**Figure S5:**
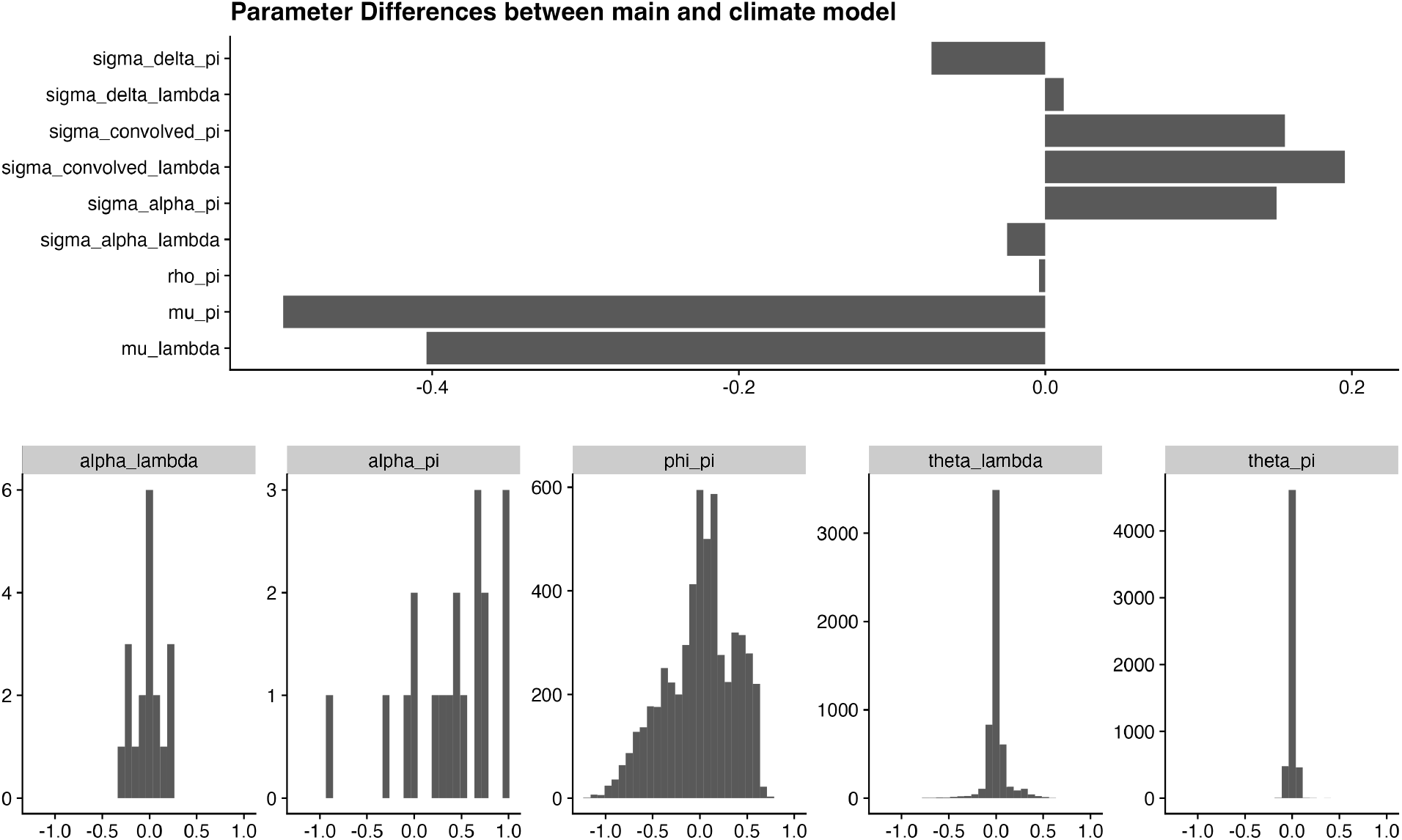
Differences in parameter estimates between the main smoothing model and the climate covariate model.

## Data Availability

All data produced in the present study are available upon reasonable request to the authors

## Footnotes

1 *Município* is translated to English as municipality, but are functionally closer to US Counties by population size, geographic size, and governance.

2 Brazil has 27 Federative Units (*unidades federativas*, abbreviated as UFs), consisting of 26 states and one federal district (Brasília). Here, we refer to all 27 UFs as states.

3 http://www.worldclim.org/

4 Related to the SAR models is the field of Spatial Econometrics (see Anselin 2003 for an overview). Spatial Econometrics uses the SAR model in a maximum likelihood regression framework. Many related specifications form the suite of Spatial Econometric models, each with their own implication about spatial correlative structure and dependence (Golgher and Voss 2016)

5 In their 2019 paper, Morris et al use a logit-normal prior for *ρ*, which has mass around either extreme, indicating that the value of *ρ* should be close to 0 or 1 and is less likely to be in the middle. However, in a 2018 case study predating the publication, the same authors use a *Beta*(1*/*2, 1*/*2) prior, which has a similar U-shape.

6 This is forgiveable as BYM—who were working in digital image restoration—were the first to demonstrate how this technique could be used in other fields, which has become a foundational technique in Bayesian Disease Mapping.

7 Only the Federal District, DF, had fewer cases, at 4 over the 19 year period, but was not chosen since that UF contains only one municpality.

8 https://mc-stan.org/docs/stan-users-guide/zero-inflated.html

9 In theory, an additional optimization of the Zero-likelihood involves use of the `log-sum-exp` trick, which provides computationally efficient evaluation of `log(a+b) = log(exp(a) + exp(b))`, but this remains unexplored at present.

